# Type and developmental timing of childhood adversity predicts psychopathology symptoms in a South African birth cohort

**DOI:** 10.1101/2025.06.18.25329880

**Authors:** Sherief Y. Eldeeb, Brooke G. McKenna, Marilyn T. Lake, Nadia Hoffman, Alexandre A. Lussier, Esther Walton, Andrew J. Simpkin, Andrew D.A.C. Smith, Heather J Zar, Dan J. Stein, Erin C. Dunn

## Abstract

**Background:** Childhood adversity is widespread globally and is one of the strongest predictors of later psychopathology. However, the differential effects of type and timing of childhood adversities on childhood psychopathology remain unclear, highlighting the need to explore which life-course hypotheses (sensitive periods, accumulation of exposure, and/or recency of exposure) best explain these associations. Of particular importance, there is a lack of research in low- and middle-income countries (LMIC), where children experience higher rates of adversity relative to children in high-income countries (HIC).

**Methods:** Participants included 787 children and their mothers from a South African birth cohort, the Drakenstein Child Health Study. Mothers reported child exposure to adversity from birth to 8 years of age across six adversity categories. We used the two-stage Structured Life Course Modeling Approach (SLCMA) to examine life-course associations between childhood adversity exposures and internalizing/externalizing symptoms measured using the Child Behavior Checklist at age 8 years.

**Results:** Maternal psychopathology, maternal adverse events, child food insecurity, and child exposure to community/domestic violence had the strongest associations with child psychopathology symptoms, with varying life-course models selected. The accumulation hypothesis best explained associations of maternal adverse events (partial *R^2^*=2.3%) and child exposure to community/domestic violence (partial *R^2^*=1.6%) with internalizing symptoms. The combined middle childhood sensitive period (age 5≤8 years) and recency hypotheses model best explained associations between maternal psychopathology and internalizing (partial *R^2^*=7.0%) or externalizing (partial *R^2^*=5.1) symptoms.

**Conclusions:** We identified that different types and timing of childhood adversity confer differential risk for childhood psychopathology symptoms in this LMIC sample. Our work has implications for strategically-timed intervention and prevention strategies to improve mental health, which may need to be specifically designed for children in LMIC.

## Introduction

Mental health disorders are a leading cause of disability globally, with prevalence estimates continuing to rise (Network, 2022; World Health Organization, 2017). Childhood adversity remains one of the strongest predictors of later psychopathology, with lifetime risk doubling for those who experience childhood adversity (Green et al., 2010; McLaughlin et al., 2012). In a global study spanning 21 countries, approximately 38% of people reported at least one form of adversity during childhood; the eradication of childhood adversity was estimated to reduce the prevalence of mental health disorders by up to 42% globally (Kessler et al., 2010).

However, optimal targets and timing for intervention remain unclear, particularly for children in low- and middle-income countries (LMIC). Gaining a clearer understanding of how the type and timing of childhood adversity impacts risk for mental health disorders is an important step towards developing effective and cost-efficient prevention and intervention efforts (Schaefer et al., 2022).

Historically, childhood adversity research has disproportionally focused on WEIRD (Western, Educated, Industrialized, Rich, and Democratic) samples, despite 88% of the world’s children residing in low- and middle-income settings (Ceccarelli et al., 2022; Henrich et al., 2010; Network, 2022). Understanding the relationships between childhood adversities and risk for mental health disorders in LMIC is crucial to expanding generalizability of results and informing the adaptation of interventions to differing cultural and socioeconomic contexts.

Children in LMIC countries, such as South Africa, are exposed to higher rates of adversity including intimate partner violence, parental substance abuse, and traumatic events (Ceccarelli et al., 2022; Stein et al., 2015). One study comparing longitudinal birth cohorts found 91% of participants from a South African cohort experienced at least one childhood adversity by age 10 years, compared to 64% and 67% in British and Dutch cohorts, respectively (Bigler et al., 2025). Further, studies examining adversities generally focus on individual-level adversities, such as interpersonal trauma, rather than community- and systemic-level factors, such as exposure to community violence and chronic food insecurity (Stirling et al., 2015). Children in LMIC are disproportionately exposed to these macro-level adversities, and increasing evidence points to their negative impact on psychopathology risk (Hyde et al., 2022).

Beyond examining the *types* of childhood adversity that influence risk for psychopathology, it is also vital to understand how *timing* of exposure may alter risk. Evidence from a recent systematic review (Schaefer et al., 2022) and empirical studies suggest both type and timing of childhood adversity influences the development of childhood psychopathology (i.e., internalizing and externalizing) symptoms, which are early indicators of later psychopathology risk (Dunn et al., 2018; Farooq et al., 2024; Juen et al., 2024). Life-course theory proposes the timing of adversity exposure may differentially impact outcomes, based on: exposure during a *sensitive period* of development, *accumulation* of exposure across childhood, and the *recency* of an exposure. Sensitive period hypotheses posit there are specific developmental periods when an exposure disproportionately impacts outcomes (Ben-Shlomo & Kuh, 2002; Frankenhuis & Fraley, 2017; Schaefer et al., 2022). Accumulation hypotheses contend that the number of exposures linearly increases risk, regardless of timing (Evans et al., 2013; Guidi et al., 2020; Slopen et al., 2018). Recency hypotheses argue adversities occurring closer to the timing of the outcome have stronger effects (Shanahan et al., 2011). Research conducted in HIC contexts, like the United Kingdom and the Netherlands, provides mixed evidence for developmental timing (Bagner et al., 2010; Yoon, 2022), accumulation (Buss et al., 2011; Hogye et al., 2022; Mäntymaa et al., 2012; Rogosch et al., 2011), and recency of exposure (Dunn et al., 2018) predicting childhood psychopathology symptoms. Very few studies have examined which life-course hypotheses best explain child psychopathology symptoms in LMIC contexts, with a single cross-sectional study conducted in Tanzania finding the recency of exposure as the strongest predictor of childhood psychopathology compared to factors such as accumulation, exposure at certain ages, and severity of exposure (Juen et al., 2024).

In the current study, we investigated which of three life-course hypotheses (sensitive period, accumulation of risk, and/or recency of exposure) best explains the associations between early adversity and childhood internalizing/externalizing symptoms. We examined these adversities in a LMIC birth cohort of South African children who are disproportionately impacted by poverty and other childhood adversities relative to children represented in HIC life-course studies (Dunn et al., 2018; Farooq et al., 2024; Moss et al., 2023). We explored a broad range of adversity types relative to previous work, with exposures spanning household-level – maternal psychopathology, substance abuse, intimate partner violence (IPV), and maternal adverse events – and the community-level – child food insecurity and exposure to community and domestic violence.

## Methods

### Participants and Procedure

Data were obtained from the Drakenstein Child Health Study (DCHS), a multidisciplinary longitudinal birth cohort from the Western Cape of South Africa (Stein et al., 2015; Zar et al., 2015). Pregnant mothers were enrolled at 20-28 weeks’ gestation from two clinics, Mbekweni and TC Newman, during routine antenatal care appointments between March 2012 and March 2015. The Mbekweni clinic predominantly serves an isiXhosa-speaking Black African population, whereas the TC Newman clinic predominantly serves an Afrikaans-speaking mixed-race population of individuals self-identifying as ‘Coloured’^1^. Both clinics are located in the peri-urban Drakenstein subdistrict. To be eligible for the study, women needed to: (1) be 18 years of age or older, (2) intend to attend antenatal care at one of the two clinics, and (3) intend to remain in the area for at least a year.

### Measures

Mothers answered in-person questionnaires on child exposure to adversity, psychopathology symptoms, and demographic information. Trained study staff administered measures in the preferred language of parents using the Research Electronic Data Capture (REDCap) platform (Harris et al., 2019; Harris et al., 2009). All study measures were collected within a target range of one month prior and three months following the child’s birthdate, except during the COVID-19 pandemic during which the range was expanded to six months following the child’s birthdate for most measures (**Figure S1**).

### Predictors: Childhood Adversity

We assessed six types of childhood adversity: (1) maternal psychopathology, (2) maternal adverse events, (3) child food insecurity, (4) child exposure to community and domestic violence, (5) intimate partner violence between mother and a partner, and (6) maternal substance abuse. Exposure to childhood adversity was measured between three to 12 timepoints, depending on category, from birth to 8 years of age (see **Table S1** for a list of all measures and administered timepoints). Measures assessing adversity exposure at birth or during the prenatal period were excluded from the analyses. Participants were dichotomously coded as exposed or unexposed to each category of adversity. Adversity measures are introduced below, with more detailed descriptions and psychometrics provided in **Appendix S1**. Each adversity was analyzed independently.

1. **Maternal Psychopathology.** Maternal psychopathology was measured using two questionnaires. The first, the *Beck Depression Inventory (BDI-II)*, is an established and validated 21-item measure assessing the severity of major depressive disorder symptoms, including sadness, pessimism, anhedonia, mood changes, and difficulty sleeping over the last two weeks (Beck et al., 1996).

The second questionnaire, the *Self-Reporting Questionnaire (SRQ-20)*, is a 20-item tool endorsed by the World Health Organization (WHO) to capture non-psychotic psychiatric symptoms, including symptoms of depressive and anxiety disorders (Harding et al., 1980).

2. **Maternal Adverse Life Events.** Maternal adverse life events were measured using the 17-item *Life Events Questionnaire (LEQ)* (Myer et al., 2008). Items are scored according to whether the event occurred in the last 12 months (0 = no, 1 = yes).

3. **Child Food Insecurity.** An adapted version of the US Department of Agriculture Short Form Household Food Security Scale (USDA-HFSS) was used to assess perceived household food insecurity resulting from financial difficulties in the last 6 months (Bickel et al., 2000).

4. **Child Exposure to Community and Domestic Violence.** Child exposure to community and domestic violence were measured using the 35-item *Child Exposure to Violence Checklist (CECV)*, a validated measure of child exposure to violence in South Africa (Bruwer et al., 2008; Fincham et al., 2009; Martin et al., 2013). Lifetime exposure was measured until 4.5 years, and past-year exposure was measured at later timepoints.

5. **Maternal Intimate Partner Violence (IPV).** Maternal intimate partner violence (IPV) was measured using the 12-item *Intimate Partner Violence (IPV) Questionnaire*, a measure adapted from the WHO country study on women’s health and domestic violence against women (Jewkes, 2002) and the Women’s Health Study in Zimbabwe (Shamu et al., 2011).

6. **Maternal Substance Abuse.** Maternal substance abuse was measured using the *Alcohol, Smoking, and Substance Involvement Screening Test (ASSIST)*. The ASSIST is a 8-item questionnaire developed by the WHO to measure substance use across 10 categories: tobacco, alcohol, cannabis, cocaine, amphetamine-type stimulants, inhalants, sedatives, hallucinogens, opioids, and other drug use over the past three months (Group, 2002). In the current study, maternal substance abuse was dichotomized such that participants with scores in the low-risk range for all substances, excluding tobacco, were considered unexposed (0) and participants with scores in the moderate- and high-risk ranges for any substance, excluding tobacco, were considered exposed (1). Maternal tobacco use was not considered an adversity exposure, but rather was included as a covariate at birth.

### Outcomes: Internalizing/Externalizing Problems

Parents reported on child psychopathology symptoms when children were 8 years old using two widely used measures: the Child Behavior Checklist for Ages 6-18 (CBCL/6-18) and the Strengths and Difficulties Questionnaire (SDQ). Our primary analyses focused on the CBCL/6-18, as it more comprehensively assesses symptomatology (via 113 items, versus 25 items in the SDQ), demonstrates stronger internal and interrater reliability metrics (Achenbach et al., 2008; Goodman, 2001), and offers greater clinical interpretability given its use of normed T-scores and defined risk ranges relative to the raw continuous scores provided by the SDQ. Nevertheless, many life-course studies solely use the SDQ. To facilitate greater comparability with these studies, we repeated all analyses using the SDQ as our secondary outcome (**Supplemental Materials**).

#### Covariates

### Sociodemographic Questionnaire

A questionnaire adapted from the SASH study was administered to collect sociodemographic information such as age, marital status, and income (Herman et al., 2009; Myer et al., 2008). All analyses adjusted for the following covariates assessed at birth: child race/ethnicity, child sex, maternal age, maternal marital status, number of mother’s previous pregnancies, highest level of maternal education, owned assets, household income, maternal smoking, and maternal HIV status. Covariates were selected for their theoretical role as potential confounders. Maternal smoking and HIV status at birth were included due to the high relative prevalence in this population, as in previous studies using the DCHS cohort (Feil et al., 2022; Moyakhe et al., 2023; Stein et al., 2015).

## Data Analysis

### Multiple Imputation

Among participants with complete outcome data (n=787), some were missing data on covariates (up to 12.8%) and/or exposure variables (0.1%-23.4%) (see comparisons of total and analytic samples in **Table S2** and **Figure S2**). To maximize our sample size, and in turn improve statistical power and reduce biased conclusions from these data, we conducted multiple imputation using the MICE package in R version 4.3.1 (van Buuren & Groothuis-Oudshoorn, 2011) within and across timepoints (**Appendix S2**).

### Quantifying Life-course Hypotheses

We defined three developmental periods (very early childhood: ≤ 3 years old, early childhood: 3≤5, middle childhood: 5≤8) for each adversity category. These age ranges are consistent with previous work and were chosen to facilitate comparisons between studies (Dunn et al., 2017; Dunn et al., 2018; Dunn et al., 2019). We considered a child as exposed to a specific adversity if they were exposed at any timepoint within each developmental period.

#### Sensitive Period

We considered three sensitive period hypotheses corresponding to the three developmental periods for each adversity, with one exception. For child exposure to community and domestic violence only two developmental periods – the first five years (≤5 years old) and middle childhood (5≤8) – were tested due to the absence of past-year exposure data prior to age 5.

#### Accumulation

Accumulation of exposure was defined as the sum of exposed developmental periods for each adversity category. Values ranged from 0 (not exposed in any developmental period) to 3 (exposed during all developmental periods).

#### Recency

Recency of exposure was defined as the weighted sum of exposed developmental periods for each adversity category, weighted by the most recent timepoint represented in the developmental period (i.e., weights of 3 for very early childhood, 5 for early childhood, and 8 for middle childhood). Thus, children exposed in more recent developmental periods had higher recency scores relative to children exposed earlier in development. For example, if a participant was exposed in the very early childhood (3) and middle childhood (8) developmental periods, their value was 11. Values ranged from 0 (not exposed in any developmental periods) to 16 (exposed during all developmental periods).

### Preliminary Analyses

Preliminary univariate descriptive statistics were used to examine the frequency of demographics, adversity exposures, and covariates. Bivariate tetrachoric correlations were conducted to examine the average correlations within each adversity category across developmental periods, and the average correlations within each developmental period across adversity categories. Multiple regression analyses were conducted to examine the “ever-exposed” relationship for each adversity category on psychopathology outcome variables.

### Primary Analyses: Structured Life-course Modeling Approach (SLCMA)

The two-stage Structured Life-course Modeling Approach (SLCMA; pronounced slick-mah; Smith et al., 2016; Smith et al., 2015; Smith et al., 2021) was utilized to determine which life-course hypotheses, individually or in combination, best explained the associations between each type of adversity exposure and symptoms of psychopathology. A full description of the two-stage process of SLCMA is described in the **Supplemental Materials**. Significance was set at a p-value of .05. We present all results, regardless of p-value, as LARS enables the selection of the best-fitting hypotheses through examination of elbow plots rather than relying on significance thresholds (Smith et al., 2021) (**Appendix S2**).

### Sensitivity Analyses

We conducted two sensitivity analyses to assess whether our findings varied by differences in how life-course models were operationalized or covariates included. First, we examined, for all adversities other than maternal psychopathology, whether results remained consistent when controlling for maternal postpartum depression, as measured via the Edinburgh Postpartum Depression Scale (EPDS; Cox et al., 1989). As in previous life-course hypothesis work (Dunn et al., 2018), maternal postpartum depression was included as a covariate to control for the potential bias maternal mood may have on reporting of child adversity exposure and psychopathology (Chilcoat & Breslau, 1997; Holt et al., 2008; Ringoot et al., 2015). Second, we tested whether averaging or summing data from individual timepoints influenced the life-course hypotheses selected, to assess whether combining measurement timepoints into developmental periods may have unduly influenced results (results reported in **Supplemental Material**).

## Results

### Sample Demographics

A total of 1225 mothers were originally enrolled in the study, with 1137 giving live birth to 1143 children. Data from 787 child-mother pairs were available after removing those with non-singleton births (n=11 children) and without complete outcome data (n=339).

The analytic sample largely comprised families from disadvantaged backgrounds. Most mothers were single (60%), completed less than secondary education (62.9%), and earned less than half the national median household income at the time of data collection (87.6%; GlobalData, 2022). Children were primarily of Black African (60.6%) descent, followed by those of Coloured backgrounds (39.2%). Male and female children were equally represented (50.3% female) (**Table 1**).

**Table 1.** Sample demographics.

|  | Mbekweni<br>(N=479) | TC Newman<br>(N=308) | Overall<br>(N=787) |
| --- | --- | --- | --- |
| **Child Ethnicity<sup>1</sup>** |  |  |  |
| Black African | 474 (99.0%) | 4 (1.3%) | 478 (60.6%) |
| Coloured | 4 (0.8%) | 304 (98.7%) | 308 (39.2%) |
| Missing | 1 (0.2%) | 0 (0%) | 1 (0.1%) |
| **Sex** |  |  |  |
| Female | 252 (52.6%) | 144 (46.8%) | 396 (50.3%) |
| Male | 227 (47.4%) | 164 (53.2%) | 391 (49.7%) |
| **Marital Status** |  |  |  |
| Single | 302 (63.0%) | 170 (55.2%) | 472 (60.0%) |
| Married/Cohabiting | 176 (36.7%) | 138 (44.8%) | 314 (39.9%) |
| Missing | 1 (0.2%) | 0 (0%) | 1 (0.1%) |
| **Highest Maternal Education at Birth** |  |  |  |
| Lower than Secondary | 302 (63.0%) | 193 (62.7%) | 495 (62.9%) |
| Secondary or Higher | 177 (37.0%) | 115 (37.3%) | 292 (37.1%) |
| **Maternal HIV at Birth** |  |  |  |
| Negative | 298 (62.2%) | 298 (96.8%) | 596 (75.7%) |
| Positive | 181 (37.8%) | 10 (3.2%) | 191 (24.3%) |
| **Number of Previous Pregnancies** |  |  |  |
| 0 | 155 (32.4%) | 114 (37.0%) | 269 (34.2%) |
| 1 | 185 (38.6%) | 107 (34.7%) | 292 (37.1%) |
| 2 | 92 (19.2%) | 55 (17.9%) | 147 (18.7%) |
| 3+ | 45 (9.4%) | 30 (9.7%) | 75 (9.5%) |
| Missing | 2 (0.4%) | 2 (0.6%) | 4 (0.5%) |
| **Maternal Age at Birth (in years)** |  |  |  |
| Mean (SD) | 27.8 (5.75) | 26.2 (5.53) | 27.2 (5.71) |
| Median [Min, Max] | 27.0 [18.0, 45.0] | 25.0 [18.0, 44.0] | 26.0 [18.0, 45.0] |
| **ASSIST Tobacco Score<sup>2</sup>** |  |  |  |

|  | **Mbekweni**<br>**(N=479)** | **TC Newman**<br>**(N=308)** | **Overall**<br>**(N=787)** |
| --- | --- | --- | --- |
| Mean (SD) | 1.13 (5.16) | 11.9 (11.4) | 5.44 (9.79) |
| Median [Min, Max] | 0 [0, 31.0] | 12.0 [0, 31.0] | 0 [0, 31.0] |
| Missing | 66 (13.8%) | 33 (10.7%) | 99 (12.6%) |
| **Asset Sum<sup>3</sup>** |  |  |  |
| Mean (SD) | 6.45 (1.99) | 7.24 (1.71) | 6.76 (1.92) |
| Median [Min, Max] | 7.00 [2.00, 11.0] | 7.00 [2.00, 10.0] | 7.00 [2.00, 11.0] |
| **Household Income<sup>4</sup>** |  |  |  |
| <R1000/m | 184 (38.4%) | 93 (30.2%) | 277 (35.2%) |
| R1000-5000/m | 249 (52.0%) | 163 (52.9%) | 412 (52.4%) |
| >R5000/m | 46 (9.6%) | 52 (16.9%) | 98 (12.5%) |

<sup>1</sup> In South Africa, the apartheid system divided individuals into different racial groups (e.g., “Black African”, “Coloured”, “White”). Under democracy these categories continue to be employed, partly for reasons of redress and partly due to South Africans’ cultural embrace and celebration of these identities (Dooms & Chutel, 2023). In DCHS, participants self-identify as “Black African” or “Coloured”. We are aware that these categories are not widely used internationally and may have painful racist connotations in other cultures. At the same time, we think it is important to honor South African participants’ identities and culture by using the language and terminology they self-report. By using these categories, we are not aiming to reify social categories but instead hope to contribute to the study of ongoing health disparities stemming from systemic racism.
<sup>2</sup> A higher ASSIST Tobacco Score indicates greater tobacco usage during the antenatal visit.
<sup>3</sup> Asset sum is determined by the endorsement of 13 items, such as access to electricity, running water, or a motor vehicle, where a higher score indicates higher assets.
<sup>4</sup> The median household income in South Africa (2015) was R11,294 per month.

### Adversity Characteristics

Adversity exposure prevalence ranged from 30.9% (child food insecurity) to 76.9% (maternal adverse events; **Table 2**). Exposure prevalence tended to be highest in the earliest developmental period measured (very early childhood), except for child food insecurity and child exposure to community and domestic violence. Adversities were moderately correlated both within adversity type across time (*r*=.22 to .53; **Figure 1**), as well as between adversities (*r*=.23 to .38; **Figure 1**).

**Figure 1.**
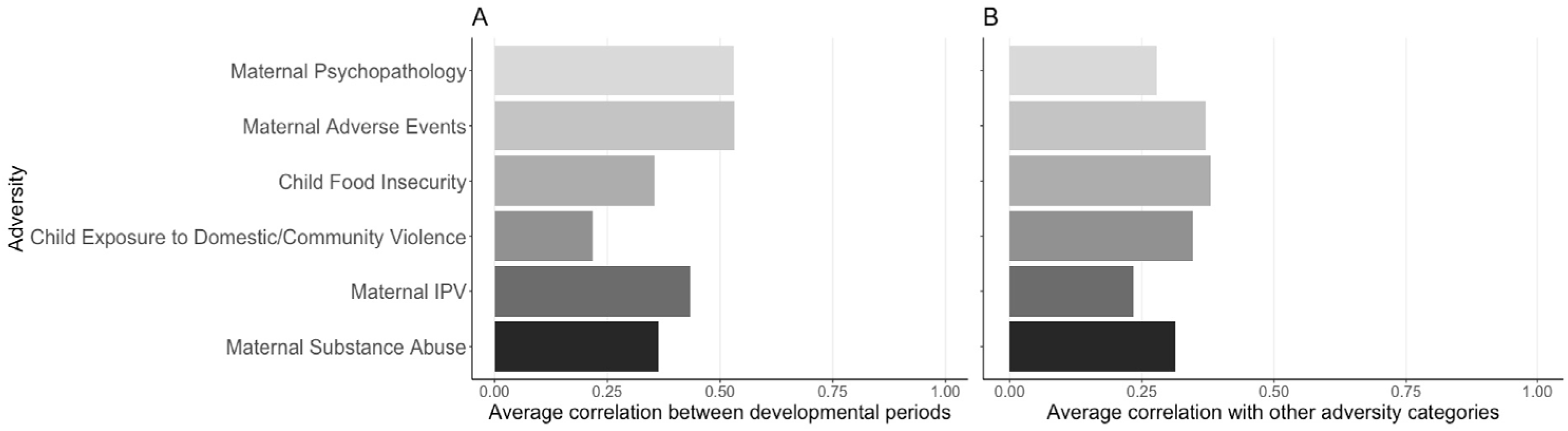
Average tetrachoric correlations between periods and adversities. **A)** average tetrachoric correlation within each adversity category across developmental periods. **B)** depicts the average tetrachoric correlation of each adversity with all other adversity categories. IPV = intimate partner violence.

**Table 2.** Adversity exposure prevalence across developmental period.

| **Adversity Category** | **Very Early Childhood**<br>**($\leq 3$)** | **Early Childhood**<br>**($3 \leq 5$)** | **Middle Childhood**<br>**($5 \leq 8$)** | **Ever Exposed** |
| --- | --- | --- | --- | --- |
| Maternal Psychopathology | 196 (24.9%) | 110 (14.0%) | 61 (7.8%) | 259 (32.9%) |
| Maternal Adverse Events | 461 (58.6%) | 313 (39.8%) | 184 (23.4%) | 605 (76.9%) |
| Child Food Insecurity | 118 (15.0%) | 93 (11.8%) | 126 (16.0%) | 243 (30.9%) |
| Child Exposure to Community and Domestic Violence* | 192 (24.4%) |  | 346 (44.0%) | 415 (52.7%) |
| Maternal Intimate Partner Violence | 303 (38.5%) | 116 (14.7%) | 61 (7.8%) | 363 (46.1) |
| Maternal Substance Abuse | 262 (33.3%) | 193 (24.5%) | 124 (15.8%) | 362 (46.0%) |

\*Note. Child Exposure to Community and Domestic Violence developmental periods are first five years (age $\leq 5$ years) and middle childhood (age $5 \leq 8$ years)

Overall exposure, irrespective of timing, was associated with increased internalizing and externalizing symptoms for all forms of childhood adversity. After controlling for covariates, maternal psychopathology explained the greatest variance in both internalizing (partial *R^2^*=3.5%) and externalizing (partial *R^2^*=2.4%) symptoms. After that, child exposure to domestic and community violence (partial *R^2^*=2.1%), child food insecurity (partial *R^2^*=1.5%), and maternal IPV (partial *R^2^*=0.9%) explained the most variance for externalizing symptoms. Maternal adverse events (partial *R^2^*=1.8%), child exposure to domestic and community violence (partial *R^2^*=1.6%), and child food insecurity (partial *R^2^*=1.5%) explained the most variance for internalizing symptoms. Unstandardized effect estimates and partial *R^2^* are shown in **Figure 2**.

**Figure 2.**
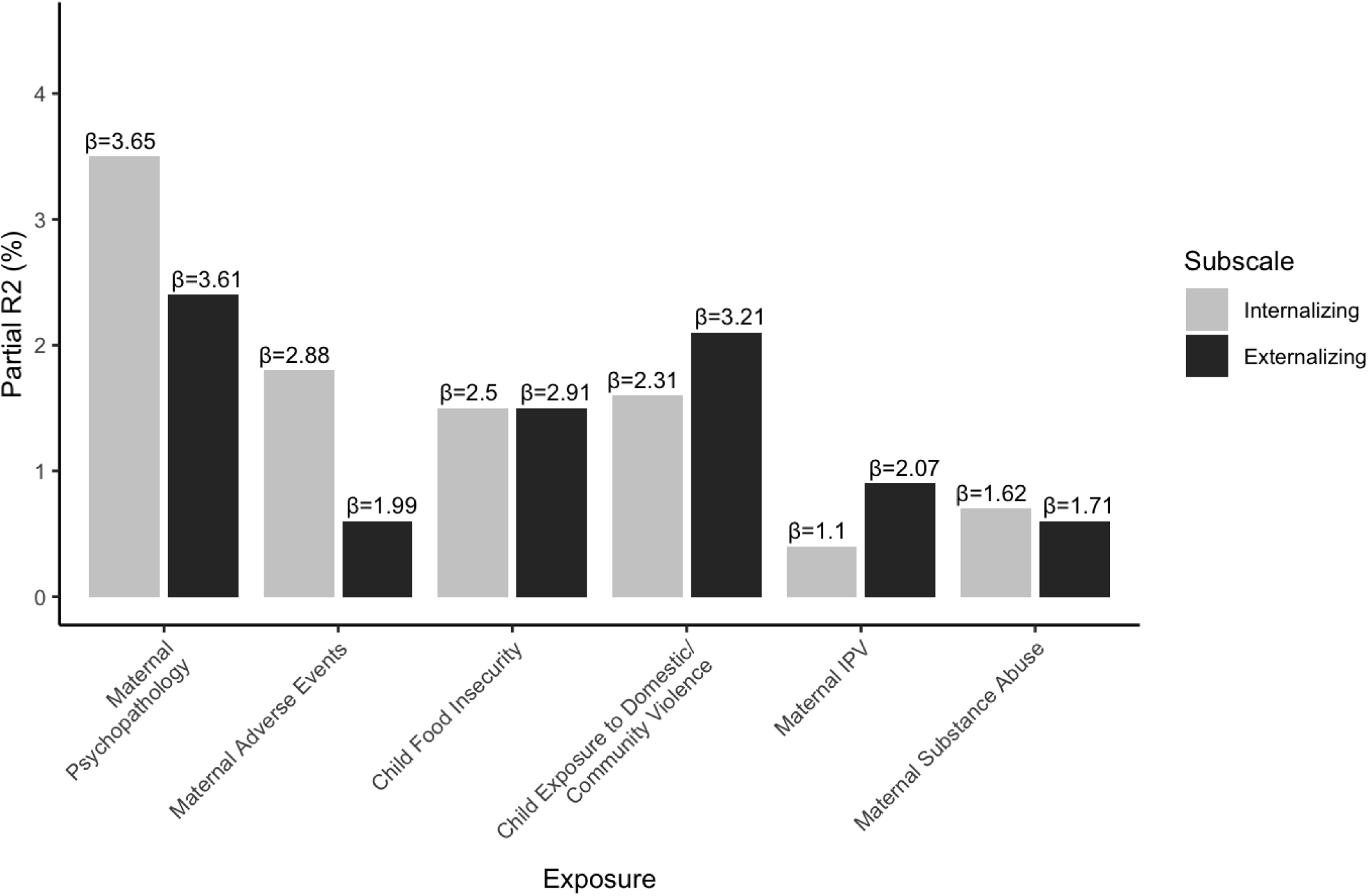
Associations of exposure to each type of adversity with CBCL-measured psychopathology symptoms. Bars display variance explained (*R^2^*) by each adversity. Values displayed above each bar depict unstandardized effect estimates. Results were obtained from ever-exposed – i.e., irrespective of life-course hypothesis selection – multiple regression analyses, adjusted for covariates. IPV = intimate partner violence. CBCL = Child Behavior Checklist.

### Life-course model selection

Results from the two-stage SLCMA are presented in **Table 3** for each adversity. Exposure to maternal psychopathology in middle childhood increased internalizing (/3=6.27, p=0.001, 95% CI [2.69, 11.98]) and externalizing (/3=6.38, p=0.004, 95% CI [1.79, 13.12]) symptoms. Exposure to maternal adverse events in all developmental periods also increased internalizing and externalizing symptoms, following an accumulation model for internalizing symptoms (/3=1.51, p<0.001, 95% CI [0.84, 2.15]) and a recency model for externalizing symptoms (/3=0.37, p<0.001, 95% CI [0.22, 0.51]). Finally, child exposure to community and domestic violence increased internalizing symptoms (/3=1.51, p=0.001, 95% CI [0.69, 2.44]) and child food insecurity in early childhood increased externalizing symptoms (/3=4.30, p=0.002, 95% CI [1.58, 6.60]). No other adversities were associated with internalizing or externalizing symptoms at the 5% significance threshold. Of note, the same hypotheses were selected when the SDQ was used to assess child symptomatology (**Table S3**; **Figure S3**).

**Table 3.** Life-course hypotheses selected by SLCMA for each childhood adversity predicting childhood psychopathology symptoms measured via the CBCL.

| Internalizing Symptoms |  |  |  |  |  |
| --- | --- | --- | --- | --- | --- |
| Adversity Category | Partial R-squared % | Model Selected | Coefficient | P-value | CI (95%) |
| Maternal Psychopathology | 7.0 | Middle Childhood | 6.27 | .001 | 2.69 - 11.98 |
|  |  | Recency | .20 | .2 | -.30 - .45 |
| Maternal Adverse Event | 2.3 | Accumulation | 1.51 | <.001 | .84 - 2.15 |
| Child Food Insecurity | 2.0 | Early Childhood | 2.8 | .06 | -.85 - 8.07 |
|  |  | Recency | .16 | .36 | -.52 - .41 |
| Child Exposure to Community and Domestic Violence | 1.6 | Accumulation | 1.61 | .001 | .69 - 2.44 |
| Maternal IPV | .6 | Middle Childhood | 1.22 | .61 | -16.3 - 7.58 |
|  |  | Recency | .13 | .30 | -.57 - 1.07 |
| Maternal Substance Abuse | .6 | Middle Childhood | .95 | .48 | -8.34 - 6.94 |
|  |  | Recency | .09 | .39 | -.56 - .70 |
| Externalizing Symptoms |  |  |  |  |  |
| Adversity Category | Partial R-squared % | Model Selected | Coefficient | P-value | CI (95%) |
| Maternal Psychopathology | 5.1 | Middle Childhood | 6.38 | .004 | 1.79 - 13.12 |
|  |  | Recency | .21 | .23 | -.38 - .51 |
| Maternal Adverse Event | 2.6 | Recency | .37 | <.001 | .22 - .51 |
| Child Food Insecurity | 1.4 | Early Childhood | 4.30 | .002 | 1.58 - 6.60 |
| Child Exposure to Community and Domestic Violence | 1.5 | Middle Childhood | .93 | .53 | -7.17 - 3.80 |
|  |  | Accumulation | 1.28 | .12 | -1.18 - 5.65 |
| Maternal IPV | .4 | Accumulation | 1.11 | .12 | -.87 - 2.1 |
| Maternal Substance Abuse | .4 | Accumulation | .96 | .13 | -.08 - 1.77 |

Note. The table presents results from 12 separate regression models, one for each adversity category and symptom outcome. All regression models adjusted for covariates. Coefficients represent unstandardized effect estimates. Bolded values indicate $p < .05$ .

Figure 3 displays effect estimates across developmental periods for life-course models demonstrating the strongest effects between childhood adversities and child psychopathology symptoms. We present all other elbow plots in **Figure S4** and life-course models in **Figure S5.**

**Figure 3.**
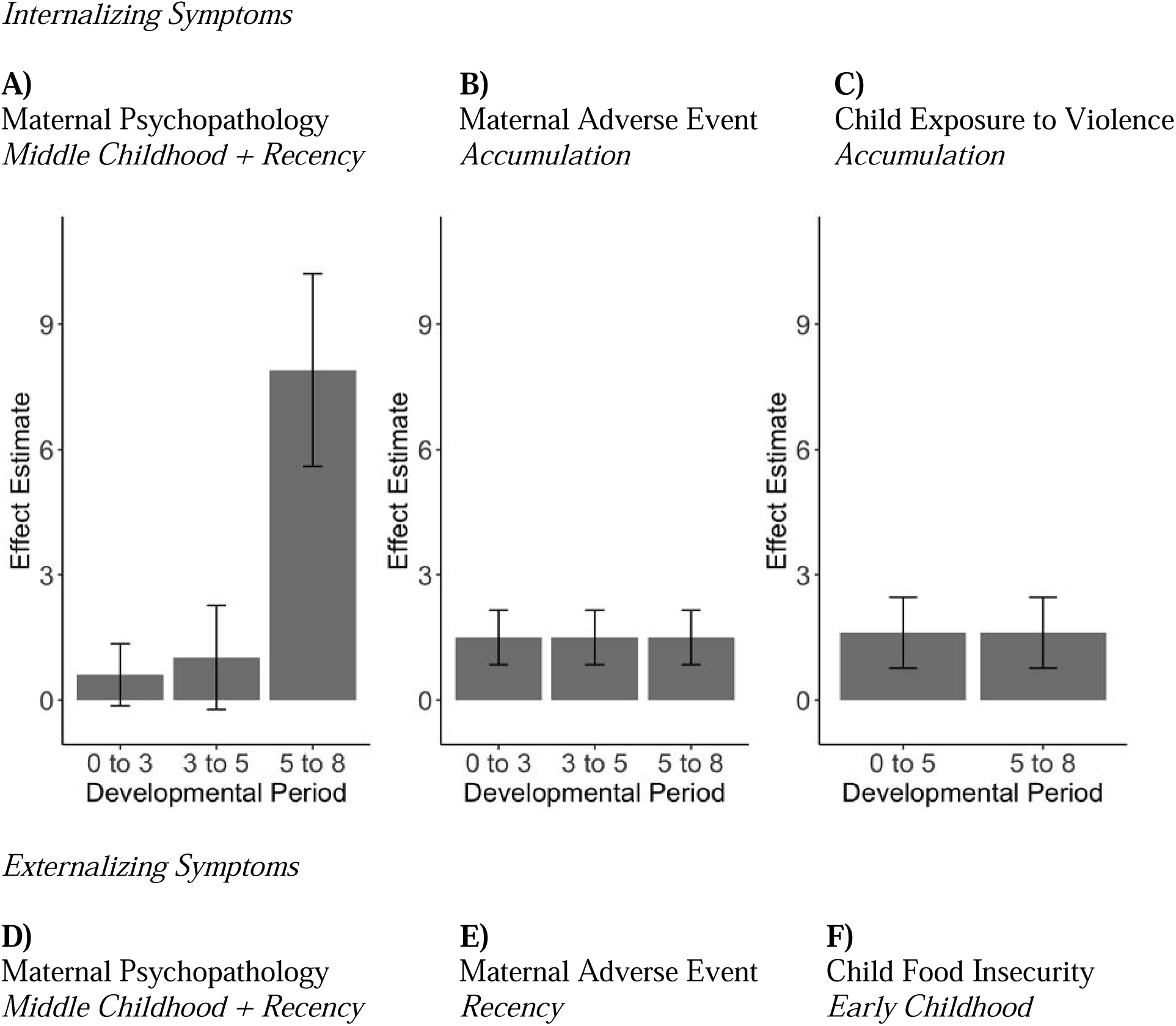
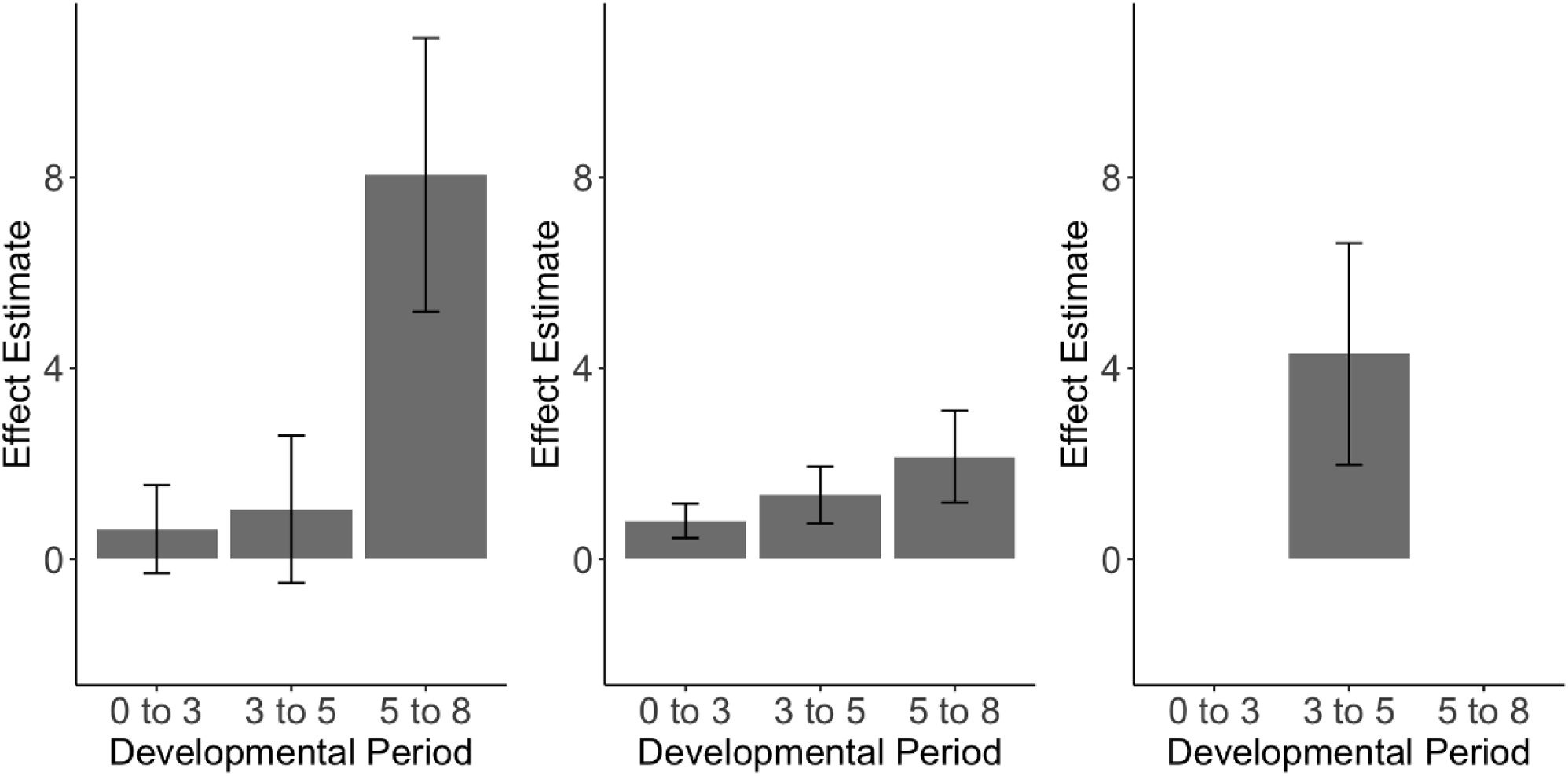
Effects of adversity exposure during each developmental period, based on the selected SLCMA life-course hypotheses via the CBCL. Figures illustrate the weighted effect of exposure during each developmental period based on the life-course hypothesis/es selected for each adversity. The y-axis was kept consistent across all figures, including negative values. *Internalizing Symptoms.* Panel A: For maternal psychopathology, the effect of exposure increases with each developmental period and there is an additional increased effect for exposures in middle childhood (age 5 ≤ 8 years). Panel B: For maternal adverse events, the effect of exposure is the same in each developmental period. Panel C: For child exposure to domestic and community violence, the effect of exposure is the same in each developmental period. *Externalizing Symptoms.* Panel D: For maternal psychopathology, the effect of exposure increases with each developmental period and there is an additional increased effect for exposures in middle childhood (age 5 ≤ 8 years). Panel E: For maternal adverse events, the effect of exposure increases with each developmental period. Panel F: For child food insecurity, exposure primarily had an effect if it occurred during the early childhood developmental period (age 3 ≤ 5 years). SLCMA = Structure Life-course Modeling Approach. CBCL = Child Behavior Checklist.

### Sensitivity Analyses

Results were largely similar when controlling for maternal postpartum depression (**Table S4**) and when utilizing alternative life-course theory operationalizations (**Table S5**).

## Discussion

Our findings suggest both the type and timing of childhood adversity influence risk for childhood psychopathology symptoms among children in a LMIC context. These differential timing and adversity effects align with previous evidence from HIC contexts (Dunn et al., 2020; Dunn et al., 2018; Farooq et al., 2024) and highlight the importance of parsing type and timing of childhood adversity exposure to inform risk evaluation, prevention, and targeted intervention approaches.

Timing was particularly important in the case of maternal psychopathology, with later exposure increasing risk. A combination of recency and the middle childhood sensitive period hypotheses were selected, indicating that risk for childhood psychopathology symptoms increased with later age of exposure and additional risk was conferred if the child was exposed during middle childhood (ages 5 to 8). The selection of recency and the middle childhood sensitive period hypotheses are somewhat inconsistent with previous evidence that earlier exposure to maternal psychopathology exerts greater risk for later psychopathology symptoms (Goodman et al., 2011; Luby et al., 2020) or that timing does not matter (Farooq et al., 2024). However, Farooq and colleagues (2024) found cumulative exposure to parental mental health problems best predicted later depression across two UK cohorts, with exposure during middle childhood conferring the greatest risk across the developmental periods measured. Together, these findings suggest middle childhood may warrant attention as a developmental period when maternal psychopathology exposure may be especially influential in shaping child psychopathology symptoms. Indeed, middle childhood is a time of rapid emotional development in children, especially in the areas of empathy and emotion regulation (Berk, 2022; De Raeymaecker & Dhar, 2022). Developmental advances in this period may therefore be more sensitive to symptoms associated with maternal psychopathology such as negative emotions, withdrawal, and low attachment security (Lovejoy et al., 2000; Suveg et al., 2011; Whittenburg et al., 2023). Future research is needed to disentangle the potential mechanisms through which maternal psychopathology during middle childhood may confer elevated risk for psychopathology symptoms.

Overall it is also worth noting that, irrespective of timing, maternal psychopathology demonstrated the strongest associations with internalizing and externalizing symptoms relative to all other childhood adversities. Maternal psychopathology may be uniquely potent compared to other adversity types due to the compound influence of both genetics and environment.

Heritability studies highlight the role of genetics in the transmission of psychopathology symptoms from parent to child, and adoption studies support the additional role of environmental exposure to maternal psychopathology symptoms (Cioffi et al., 2021; Jami et al., 2020).

Maternal psychopathology may also have a particularly profound influence on child symptoms due to their bidirectional, or transactional, nature whereby parent and child symptoms influence one another across time (Buchanan-Pascall et al., 2017). This finding highlights maternal psychopathology as an important intervention target to reduce child risk for psychopathology, especially in populations such as within the DCHS cohort where mothers are predominantly the sole caretakers of children. Indeed, prior studies examining parent interventions aimed at reducing maternal psychopathology indicate small to moderate effects in improving child symptoms (Cuijpers et al., 2015; Gunlicks & Weissman, 2008), including in LMIC contexts (Hoffmann et al., 2022). Of note, many of these interventions target mothers during their child’s first years of life (Cuijpers et al., 2015). Our work suggests interventions attempting to reduce maternal psychopathology and improve child symptoms may have greater success if implemented when children are between ages 5 and 8.

By contrast, food insecurity exposure during the early childhood sensitive period (ages 3 to 5) best predicted childhood psychopathology symptoms. Our results in this LMIC study differ from those conducted in HIC, which have not pointed to a sensitive period when exposure is disproportionately associated with impaired behavioral and emotional development (Shankar et al., 2017). Our findings raise important questions about why the early childhood sensitive period in particular emerged in this LMIC context. One explanation may be that food insecurity serves as a proxy for malnutrition. Indeed, proper nutrition plays a particularly important role for neural plasticity prior to age 5 (Georgieff et al., 2015) and the pre-frontal cortex, an area crucial for emotion regulation, displays rapid development from ages 3 to 5 (Hodel, 2018). Given that measures of deprivation, such as poverty and food insecurity, have been linked with slowed development of the pre-frontal cortex (Hodel, 2018), it is possible that food insecurity between ages 3 to 5 disproportionately impacts child pre-frontal cortex development, which, in turn, influences later development of psychopathology symptoms. Taken together, our findings suggest future research would benefit from better understanding how timing differences influence the impact of food insecurity on child psychopathology symptoms.

Specificity in the timing of exposure mattered less for maternal adverse events and child exposure to domestic/community violence, where the accumulation of exposures was most often predictive of child symptoms. These findings align with previous research on the importance of cumulative life stressors for both parents and children in predicting childhood psychopathology symptoms (Benito-Gomez et al., 2019; Evans et al., 2013; Hogye et al., 2022). Research conducted in the US in both cross-sectional and prospective longitudinal samples support our findings, with additional exposure of community violence or maternal stressful life events conferring corresponding increased risk for child internalizing and/or externalizing symptoms (Fleckman et al., 2016; Rudd et al., 2022). By contrast, recency of child abuse was the best predictor of child internalizing and externalizing symptoms in a LMIC sample from Southern Tanzania, although the study was cross-sectional and retrospective which may account for the difference with our results (Juen et al., 2024). The selection of the accumulation hypothesis for maternal adverse events and child exposure to domestic/community violence suggests a more general prevention strategy may be more advantageous than targeting specific developmental periods. Overall, our study highlights the benefits of a secondary screening prevention strategy for both parents and children experiencing cumulative life stressors where additional events may indicate additional need for intervention. A risk strategy is especially critical in the context of LMIC settings where stressors are more prevalent (Bigler et al., 2025).

### Strengths and Limitations

Our study has several notable strengths. We separately examined six forms of childhood adversity in a LMIC where children are disproportionately exposed to early life adversity, focusing not only on individual-level factors, but also community- and systemic-level factors, such as food insecurity and child exposure to domestic/community violence. Despite increasing evidence that community- and systemic-level factors are negatively associated with child mental health (Fleckman et al., 2016; Gard et al., 2021), these factors remain understudied in life-course research examining risk for child psychopathology symptoms. In particular, literature examining food insecurity in LMIC has predominantly focused on cognitive and motor skill development rather than on indicators of child psychopathology symptoms (De Oliveira et al., 2021).

Moreover, by examining adversities separately rather than grouping them into a total exposure score, we could identify unique timing effects for each category of adversity. The prospective longitudinal cohort design allowed us to study timing effects with additional rigor and draw wider conclusions than cross-sectional or retrospective designs.

Our study also had limitations. Our study had a smaller sample size compared to previous life-course work in HIC samples. Thus, we grouped exposures into sensitive periods (e.g., early childhood, ages 1-3) rather than examining each individual timepoint. While this strategy improved statistical power, it also reduced the granularity of our findings related to timing of exposure. During the COVID-19 pandemic, the study team expanded target windows for data collection between June 10, 2021 and November 3, 2022. Although this approach increased participation, it also increased variability in child age at collection, which may have impacted the specificity of our timing findings.

## Conclusion

To our knowledge, we have conducted the first prospective study in a LMIC sample examining which life-course hypotheses best explain the associations between childhood adversities and psychopathology symptoms in middle childhood. Our findings reinforce the importance of considering the type and timing of adversity when assessing risk for child psychopathology in highly adversity-exposed populations. If replicated, our findings can inform the administration of strategically-timed interventions, as children exposed to adversity during specific developmental periods may also be more sensitive to related interventions. The WHO estimates a roughly four-fold return of global investment in anxiety and depression treatments (Chisholm et al., 2016). By incorporating optimally-timed interventions that buffer the deleterious impacts of adversity, there is potential to further maximize these benefits, particularly among children in LMIC who are disproportionately impacted by childhood adversity.

### Key Points

- Childhood adversity is widespread and is a strong predictor of later psychopathology.
- Much of the research in childhood adversity and life-course hypotheses occurs in high-income countries.
- Our work, conducted in the low-and middle-income country (LMIC) of South Africa, found that middle childhood and recency life-course hypotheses had the strongest associations with later childhood internalizing and externalizing symptoms.
- Future research should continue exploring sensitive periods and mechanisms of childhood adversity in LMIC to inform prevention and intervention efforts.

## Abbreviations

(LMIC): low- and middle-income countries
(DCHS): Drakenstein Child Health Study
Structured Life-course Modeling Approach

## Data Availability Statement

The Drakenstein Child Health Study is committed to the principle of data sharing. De-identified data will be made available to requesting researchers as appropriate. Requests for collaborations are welcome. More information can be found on our website [http://www.paediatrics.uct.ac.za/scah/dclhs].

## Ethical Information

The DCHS was approved by the Faculty of Health Sciences, Human Research Ethics Committee, University of Cape Town (401/2009) and by the Western Cape Provincial Health Research committee. Mothers provided informed consent at enrolment and were re-consented annually. Consent was done in mother’s preferred language: English, Afrikaans or isiXhosa. Assent was obtained annually from children from 7 years of age depending on their neurocognitive ability.

## Acknowledgments

We greatly thank the parents and children who participated in this study. We would also like to thank the study staff in Paarl, the study data and laboratory teams, and the clinical and administrative staff of the Western Cape Government Health Department at Paarl Hospital and at the clinics for support of the study. Additionally, we would like to thank Alison Hoffnagle, Samantha Stoll, Isabel Schuurmans, Anke Hüls, and Anna Ruehlmann for their assistance in preparing this manuscript. This manuscript reflects the views of the authors and may not reflect the opinions or views of the NIH.

## Footnotes

1 In South Africa, the apartheid system divided individuals into different racial groups (e.g., “Black African”, “Coloured”, “White”). Under democracy these categories continue to be employed, partly for reasons of redress and partly due to South Africans’ cultural embrace and celebration of these identities Dooms, T., & Chutel, L. E. (2023). *Coloured: How Classification Became Culture*. Jonathan Ball Publishers. In DCHS, participants self-identify as “Black African” or “Coloured”. We are aware that these categories are not widely used internationally and may have painful racist connotations in other cultures. At the same time, we think it is important to honor South African participants’ identities and culture by using the language and terminology they self-report. By using these categories, we are not aiming to reify social categories but instead hope to contribute to the study of ongoing health disparities stemming from systemic racism.

