## Supplemental Materials for "Type and developmental timing of childhood adversity predicts psychopathology symptoms in a South African birth cohort"

*Appendix S1. Measures*

**Maternal Psychopathology**

Beck Depression Inventory (BDI-II)

Response options range from 0 (no experience or impairment) to 3 (constant and/or impaired experience). A total score is obtained by summing individual item responses, where a higher score is indicative of more severe depressive symptoms. The BDI-II has been widely used in studies in South Africa and has demonstrated high validity (*r*=.67-.82) and internal reliability (Cronbach’s α=.84-.93) in psychiatric and non-psychiatric populations (Makhubela & Mashegoane, 2016; Steele & Edwards, 2008; Subica et al., 2014).

Self-Reporting Questionnaire (SRQ-20)

Each item was coded based on whether the symptom was present (1) or absent (0) at the time of collection. Individual items were summed to generate a total score, with higher scores indicating greater psychological distress. The SRQ-20 has been used in other studies in South Africa and demonstrates fair discriminate validity (AUC = .74) and strong internal reliability (Cronbach’s α=.86) in the DCHS cohort (van der Westhuizen et al., 2018; van der Westhuizen et al., 2016b).

Edinburgh Postnatal Depression Rating Scale (EPDS)

The EPDS is a 10-item measure of mothers’ postnatal depressive symptoms and has been validated for measuring depression across the perinatal period (Cox et al., 1987; Westhuizen et al., 2016). At the antenatal visit, mothers reported how frequently they experienced 10 symptoms including anhedonia, difficulty sleeping, and thoughts of self-harm. Response options ranged from 0 (“Not at all”) to 3 (“All the time”). A total score is obtained by summing individual item responses, where a higher score suggests more severe depressive symptoms. The EPDS has demonstrated strong sensitivity (81%) and specificity (88%) in international studies (Levis et al., 2020).

Children were coded as exposed to maternal psychopathology if their mothers reported high levels of depressive symptoms or psychological distress on either the BDI-II (defined as total scores ≥20**)** or SRQ-20 (defined as total scores ≥8). These cut-points are consistent with prior work in the DCHS (Brittain et al., 2015; MacGinty et al., 2020; van der Westhuizen et al., 2018) and other studies (Beusenberg et al., 1994; Hanlon et al., 2015; Harpham et al., 2003). The EPDS was used for sensitivity analyses.

**Maternal Adverse Life Events**

Life Events Questionnaire (LEQ)

The LEQ includes a list of items representing life stressors and traumas mothers may have experienced within the last 12 months: serious illness/injury; physical attack; robbery; valuables stolen; death of loved one; separation from partner or spouse; break up of other close relationships; being fired from a job; retiring from a job prematurely against your will; losing job for another reason; unsuccessfully searching for a new job for more than one month; major financial crisis; problems with police; serious illness or injury to someone close to you; serious ongoing disagreements with family, friends, or anyone at work.

At the start of the study, the LEQ consisted of 17 items but was later expanded to 51 items at the 3-year timepoint. To achieve consistency of measurement over the life-course, we used only the 17 items collected at every timepoint.

In lieu of an established cut-off score, we dichotomized values according to a tertile-based cutoff score of 1. To establish this cutoff, we calculated the upper tertile (33%) of total scores at each timepoint, averaged across all timepoints, and rounded to the nearest integer. As such, mothers were considered exposed (1) if they had one or more adverse events, and unexposed (0) if they had no adverse events.

**Child Food Insecurity**

US Department of Agriculture Short Form Household Food Security Scale (USDA-HFSS)

Mothers responded Yes (1) or No (0) to five items inquiring whether meals were made smaller for children, children skipped any meals, were hungry, or went a full day without food because they could not afford more food. Items were summed for a total food insecurity score that ranged from 0 to 5. An established cut-off of 2 was used to dichotomize participants as exposed (1) or unexposed (0).

**Child Exposure to Community and Domestic Violence**

Child Exposure to Violence Checklist (CECV)

The CECV uniquely captures *child* exposure to domestic and community violence. This differs from the Intimate Partner Violence Questionnaire, which also measures aspects of domestic violence but focuses on maternal experiences. Additionally, the CECV provides a broader measure of both direct and indirect community violence outside of the home. Parents reported the frequency of their child’s exposure to multiple types of violence, including community-based crime (assault, stabbings, robbery, and shootings), domestic violence, or physical and sexual abuse. Responses ranged from 0 (“Never”) to 3 (“Often”).

Item responses were used to calculate four subscales of the CECV: witnessing community violence, community violence victimization, witnessing domestic violence, and domestic victimization. Presence or absence of exposure was based on the concept of polyvictimization, which was operationalized in a former DCHS project as a score of 1 or more on two or more CECV subscales (Tsunga et al., 2023). Therefore, scores were dichotomized such that exposure on two or more CECV subscales was considered exposed (1) and exposure to one or fewer forms was considered unexposed (0).

**Maternal Intimate Partner Violence (IPV)**

Intimate Partner Violence (IPV) Questionnaire

Mothers reported how frequently they experienced emotional, physical, and sexual IPV throughout their lifetime and in the past year. Item responses were based on a 4-point frequency scale ranging from 1 (“Never”) to 4 (“Many times”). The IPV questionnaire has strong internal consistency reliability (Cronbach’s α=.91) in the DCHS cohort (Barnett et al., 2022).

Previous studies have established scoring and dichotomization guidelines for this measure based on prior work in South Africa (Barnett et al., 2018; Dunkle et al., 2004). Scores were dichotomized such that exposure in the last year to two or more occurrences of IPV on any subscale (emotional, physical, or sexual) was considered exposed (1) and exposure to one or fewer occurrences was considered unexposed (0).

**Maternal Substance Abuse**

Alcohol, Smoking, and Substance Involvement Screening Test (ASSIST)

Prior studies indicate that scores of 0-10 for alcohol and 0-3 for all other substances are considered low risk, scores of 11-26 for alcohol and 4-26 for all other substances indicate moderate risk, and scores >26 suggest high risk of experiencing severe problems as a result of current use and dependence (Humeniuk et al., 2010). The ASSIST has demonstrated good internal consistency (Cronbach’s α=.81-.95) and sensitivity/specificity (60-93%) across studies and in a South African context (McNeely et al., 2014; van der Westhuizen et al., 2016a).

**Internalizing/Externalizing Problems**

Strengths and Difficulties Questionnaire (SDQ)

The SDQ is a 25-item measure of behavioral and emotional problems in children and adolescents in the last six months (Goodman, 1997). Response options range from 0 (“Not True”) to 2 (“Certainly True”). There are five subscales: emotional symptoms, conduct problems, hyperactivity/inattentive, peer relationships problem, and prosocial behaviors. These subscales (excluding the prosocial subscale) can be combined to form internalizing (emotional problems and peer problems) and externalizing (conduct and hyperactivity) subscales. Scores are obtained by summing individual item responses, where a higher score is indicative of more behavioral and emotional problems. The SDQ strongly correlates with the CBCL as well as clinician diagnoses of child psychopathology (Goodman & Scott, 1999; Mathai et al., 2004). The measure is widely used in African populations and has shown strong internal reliability (Cronbach’s α=.81) when utilized properly in this context (Hoosen et al., 2018; Kashala et al., 2005).

Child Behavior Checklist for Ages 6-18 (CBCL/6-18)

The CBCL/6-18 is a well-established measure of emotional and behavior problems indicative of psychopathology in youth (Achenbach & Rescorla, 2001). The 113-item scale references behaviors over the last 6 months, with responses ranging from 0 (“Not true (as far as you know)”) to 2 (“Very true or often true”). The CBCL includes nine subscales: anxiety, depression, somatic complaints, social problems, thought problems, attention problems, rule-breaking behavior, aggressive behavior, and other problems (such as problems with sleep). These subscales can be combined to form internalizing and externalizing subscales. Total scores for each subscale are obtained by summing individual item responses and converting summed scores to norm-referenced T-scores (*M* = 50, *SD* = 10) for comparability across versions of the CBCL/6-18. Higher T-scores represent greater behavioral and emotional problems, with T-scores ≥ 64 indicating ‘clinically significance’. The CBCL demonstrates strong internal consistency and reliability (Cronbach’s α=.78-.94) across internalizing and externalizing subscales in South African populations (Achenbach & Rescorla, 2001; Zieff et al., 2022).

*Appendix S2. Analyses*

**Multiple Imputation**

All exposure, covariate, and outcome variables were included in the imputation model as well as auxiliary variables including the MINI International Neuropsychiatric Interview Major Depressive Episode (Lecrubier et al., 1997; Sheehan et al., 1998), the Edinburgh Postnatal Depression Scale (EPDS; Cox et al., 1987), and the Survey of Exposure to Community Violence (SECV; Richters & Saltzman, 1990), which were included due to their relevance to imputed predictor variables (e.g., maternal psychopathology (BDI-II), child exposure to domestic and community violence (CECV) (Mainzer et al., 2024)).

As mentioned in the Methods, our measure of maternal psychopathology combined scores from the BDI-II and SRQ-20. Given that the BDI-II was not administered to participants at the 6-10 week and 6-month timepoints, BDI-II total scores were imputed at those two timepoints. In the case of the LEQ and CECV, data were considered missing and were therefore imputed if participants did not indicate whether an endorsed event had occurred within the past year. Otherwise, data for all variables were imputed at the total score level for all missing items. Five imputed datasets with 20 iterations were created and analyses were pooled following Rubin’s rule across all imputed datasets when reporting results (Rubin, 1987).

**Structured Life-course Modeling Approach (SLCMA)**

SLCMA is a two-stage process. In the first stage, we entered all life-course hypotheses into the LARS procedure to determine the life-course hypothesis (or combination of hypotheses) that explained the most variance in the outcome. Elbow plots, which depict changes in the amount of outcome variance explained at each step of the LARS procedure, measured as R^2^, were visually examined to identify the regression model to carry forward to the second step of the analysis. In the second stage, we used post-selective inference to estimate the effect estimates and confidence intervals of the selected regression model (Tibshirani et al., 2016). We adjusted for covariates using partitioned regression (i.e., Frisch-Waugh-Lovell theorem), which improves power to detect differences between groups in penalized regression analyses (Yamada, 2017; Zhu et al., 2021).

SLCMA uses Least Angle Regression (LARS; Efron et al., 2004), a model selection algorithm useful for fitting parsimonious linear regressions, to compare the competing life-course hypotheses concurrently and with greater statistical power than forward selection or multiple regression (Smith et al., 2015). Further, when used with additional post-selection inference methods, LARS is able to produce unbiased effect estimates (Lockhart et al., 2014). The SLCMA has been used to study competing life-course hypotheses in studies by our research group (Dunn et al., 2023; Dunn et al., 2018; Dunn et al., 2019) and others (Collin et al., 2015; Evans et al., 2012; Moss et al., 2023). While prior life-course studies have stratified analyses by sex to account for potential sex differences, we elected to analyze all participants together while adjusting for sex as a covariate due to the smaller sample size of our cohort (Dunn et al., 2023; Dunn et al., 2018; Moss et al., 2023).

**Alternative Life-Course Hypotheses**

**Sensitive Period.** Rather than an ever-exposed model, the average number of exposed timepoints within a developmental period was used for each adversity category. For example, in the LEQ, there are four possible timepoints of exposure in the very early childhood developmental period. If an individual were exposed in two of the four timepoints, their score would be: $\frac{1}{2}= \frac{(1+1+ 0+0)}{4}$ .

**Accumulation.** Accumulation of exposure was defined as the sum of exposed individual timepoints for each adversity category. In the LEQ, there were eight timepoints measured. Therefore, values would range from 0 (not exposed at any timepoint) to 8 (exposed during all timepoints).

**Recency.** Recency of exposure was defined as the weighted sum of exposed individual timepoints for each adversity category, weighted by the timepoint in years. In the LEQ, measurements were collected at 1, 1.5, 2, 3, 4, 5, 6, and 8 years. Therefore, values would range from 0 (not exposed at any timepoint) to 30.5 (exposed during every timepoint; 1+1.5+2+3+4+5+6+8).

*Supplemental Tables and Figures*

Table S1. Measurement of adversity exposures

| **Adversity Category** | **Level** | **Questionnaire** | **Timepoints*** | **Reference Timeframe** |
| --- | --- | --- | --- | --- |
| Maternal Psychopathology | Household | BDI | 6-10 weeks^1^, 0.5^1^, 1, 1.5, 2, 3, 4, 5, 6, 8 | Last 2 weeks |
|  |  | SRQ | 6-10 weeks, 0.5, 1, 1.5, 2, 3, 4, 5, 6, 8 | Current |
| Maternal Adverse Events | Household | LEQ | 1, 1.5, 2, 3, 4, 5, 6, 8 | Last 12 months |
| Child Food Insecurity | Community | USDA-HFSS | 1, 1.5, 2, 2.5, 3, 3.5, 4, 4.5, 5, 5.5, 6, 6.5 | Last 6 months |
| Child Exposure to Community and Domestic Violence | Community | CECV | 4.5, 6, 8 | Lifetime (54 months);  Last 12 months |
| Maternal Intimate Partner Violence | Household | IPV | 0.5, 1, 1.5, 2, 3, 4, 5, 6, 8 | Last 12 months |
| Maternal Substance Abuse | Household | ASSIST | 0.5, 1, 1.5, 2, 3, 4, 5, 6, 7 | Last 3 months |

*Note. Timepoints are in child’s years of age unless otherwise noted

^1^Timepoints fully imputed that were not administered by the DCHS team

Table S2. Comparison of sample demographics between the analytic and total sample

|  | **Analytic Sample**  **(N = 787)** | **Total Sample  (N=1137)** | **Total vs. Analytic Sample^1^** |
| --- | --- | --- | --- |
| **Child Ethnicity**^2^ |  |  |  |
| Black African | 478 (60.6%) | 626 (55.1%) | *p* = .01 |
| Coloured | 308 (39.2%) | 510 (44.9%) |  |
| Missing | 1 (0.1%) | 1 (0.1%) |  |
| **Sex** |  |  |  |
| Female | 396 (50.3%) | 554 (48.7%) | *p* = .52 |
| Male | 391 (49.7%) | 583 (51.3%) |  |
| **Marital Status** |  |  |  |
| Single | 472 (60.0%) | 678 (59.6%) |  |
| Married/Cohabiting | 314 (39.9%) | 458 (40.3%) | *p* = .91 |
| Missing | 1 (0.1%) | 1 (0.1%) |  |
| **Highest Maternal Education at Birth** |  |  |  |
| Lower than Secondary | 495 (62.9%) | 692 (60.9%) | *p* = .39 |
| Secondary or Higher | 292 (37.1%) | 445 (39.1%) |  |
| **Maternal HIV at Birth** |  |  |  |
| Negative | 596 (75.7%) | 893 (78.5%) | *p* = .16 |
| Positive | 191 (24.3%) | 244 (21.5%) |  |
| **Number of Previous Pregnancies** |  |  |  |
| 0 | 269 (34.2%) | 417 (36.7%) | *p* = .70 |
| 1 | 292 (37.1%) | 405 (35.6%) |  |
| 2 | 147 (18.7%) | 200 (17.6%) |  |
| 3+ | 75 (9.5%) | 110 (9.7%) |  |
| Missing | 4 (0.5%) | 5 (0.4%) |  |
| **Maternal Age at Birth (in years)** |  |  |  |
| Mean (SD) | 27.2 (5.71) | 26.9 (5.71) | *p* < .001 |
| Median [Min, Max] | 26.0 [18.0, 45.0] | 26.0 [18.0, 45.0] |  |
| **ASSIST Tobacco Score^3^** |  |  |  |
| Mean (SD) | 5.44 (9.79) | 6.17 (10.2) | *p* < .001 |
| Median [Min, Max] | 0 [0, 31.0] | 0 [0, 31.0] |  |
| Missing | 99 (12.6%) | 148 (13.0%) |  |
| **Asset Sum**^4^ |  |  |  |
| Mean (SD) | 6.76 (1.92) | 6.84 (2.01) | *p* < .001 |
| Median [Min, Max] | 7.00 [2.00, 11.0] | 7.00 [0, 12.0] |  |
| **Household Income**^5^ |  |  |  |
| <R1000/m >R5000/m | 277 (35.2%) | 383 (33.9%) | *p* = .59 |
| R1000-5000/m | 412 (52.4%) | 592 (52.1%) |  |
| >R5000/m | 98 (12.5%) | 159 (14.0%) |  |
| Missing | 0 (0.0%) | 1 (0.1%) |  |

*Note. A higher ASSIST Tobacco Score at birth indicates greater tobacco usage. Asset sum is determined by the endorsement of 13 items, such as access to electricity, running water, or a motor vehicle, where a higher score indicates higher assets. The median household income in South Africa (2015) was R11,294 per month.

^1^ Independent sample t-tests and chi-square tests were used to test sample equivalence.

^2^ In South Africa, the apartheid system divided individuals into different racial groups (e.g., “Black African”, “Coloured”, “White”). Under democracy these categories continue to be employed, partly for reasons of redress and partly due to South Africans’ cultural embrace and celebration of these identities (Dooms & Chutel, 2023). In DCHS, participants self-identify as “Black African” or “Coloured”. We are aware that these categories are not widely used internationally and may have painful racist connotations in other cultures. At the same time, we think it is important to honor South African participants’ identities and culture by using the language and terminology they self-report. By using these categories, we are not aiming to reify social categories but instead hope to contribute to the study of ongoing health disparities stemming from systemic racism.

^3^ A higher ASSIST Tobacco Score indicates greater tobacco usage during the antenatal visit.

^4^ Asset sum is determined by the endorsement of 13 items, such as access to electricity, running water, or a motor vehicle, where a higher score indicates higher assets.

^5^ The median household income in South Africa (2015) was R11,294 per month.

Table S3. Life-course hypotheses selected by SLCMA for childhood psychopathology symptoms measured via the SDQ

| **Internalizing Symptoms** | | | | | | |
| --- | --- | --- | --- | --- | --- | --- |
| **Adversity Category** | **Partial R-squared %** | **Model Selected** | **Coefficient** | **P-value** | **CI (95%)** | **Concordance with CBCL** |
| **Maternal Psychopathology** | 2.7 | Middle Childhood | 1.57 | **.001** | .96 – 2.15 | + |
| **Maternal Adverse Event** | .7 | Accumulation | .25 | **.03** | -.02 - .42 | + |
| **Child Food Insecurity** | .9 | Middle Childhood | .63 | **.01** | .09 – 1.07 | - |
| **Child Exposure to Community and Domestic Violence** | .7 | First Five Years | .49 | **.04** | -.08 - .85 | - |
| **Maternal IPV** | .7 | Middle Childhood | .71 | **.03** | -.01 – 1.29 | + |
| Maternal Substance Abuse | .3 | Early Childhood | .31 | .12 | -.24 - .68 | - |
| **Externalizing Symptoms** | | | | | | |
| **Adversity Category** | **Partial R-squared %** | **Model Selected** | **Coefficient** | **P-value** | **CI (95%)** | **Concordance with CBCL** |
| Maternal Psychopathology | 1.5 | Middle Childhood | .69 | .38 | -2.83 – 2.88 | + |
|  |  | Recency | .06 | .26 | -.14 - .26 |  |
| Maternal Adverse  Event | 1.5 | Early Childhood | .24 | .33 | -1.53 – 2.68 | + |
|  |  | Accumulation | .06 | .80 | -9.83 – 1.91 |  |
|  |  | Recency | .04 | .18 | -.32 – 1.61 |  |
| **Child Food Insecurity** | 1.1 | Recency | .09 | **.005** | .03 - .15 | - |
| **Child Exposure to**  **Community and**  **Domestic Violence** | 1.7 | Middle Childhood | 1.05 | **.001** | .49 – 1.45 | + |
| Maternal IPV | .5 | Recency | .07 | .11 | -.05 - .12 | - |
| Maternal Substance  Abuse | .4 | Early Childhood | .39 | .12 | -.32 - .88 | - |
|  |  | Middle Childhood | -.26 | .40 | -.81 – 1.12 |  |

Note. The table presents results from 12 separate regression models, one for each adversity category and symptom outcome. All regression models adjusted for covariates. Coefficients represent unstandardized effect estimates. Bolded values indicate p < .05. Plus (+) symbol indicates at least one common model selected in the case of a combined hypothesis with main analyses. Minus (-) symbol indicates no common model.

Table S4. Sensitivity analysis: Life-course hypotheses selected by SLCMA for childhood psychopathology symptoms measured via the CBCL, adjusting for maternal postpartum depression

| **Internalizing Symptoms** | | | | | | |
| --- | --- | --- | --- | --- | --- | --- |
| **Adversity Category** | **Partial R-squared %** | **Model Selected** | **Coefficient** | **P-value** | **CI (95%)** | **Concordance with CBCL** |
| **Maternal Psychopathology** | 6.1 | Middle Childhood | 6.48 | **<.001** | 3.10 - 16.5 | + |
|  |  | Recency | .14 | .48 | -.77 - .37 |  |
| **Maternal Adverse Event** | 2.3 | Accumulation | 1.40 | **<.001** | .71 - 2.04 | + |
| **Child Food Insecurity** | 1.7 | Early Childhood | 2.83 | **.04** | -.58 - 11.74 | + |
|  |  | Recency | .12 | .60 | -1.07 - .35 |  |
| **Child Exposure to Community and Domestic Violence** | 1.2 | Recency | .21 | .46 | -.84 - .31 | - |
| Maternal IPV | .5 | Middle Childhood | 1.47 | .33 | -7.51 - 12.00 | + |
|  |  | Recency | .08 | .58 | -1.03 - .56 |  |
| Maternal Substance Abuse | .6 | Middle Childhood | .94 | .45 | -7.83 - 7.49 | + |
|  |  | Recency | .08 | .43 | -.63 - .66 |  |
| **Externalizing Symptoms** | | | | | | |
| **Adversity Category** | **Partial R-squared %** | **Model Selected** | **Coefficient** | **P-value** | **CI (95%)** | **Concordance with CBCL** |
| **Maternal Psychopathology** | 4.3 | Middle Childhood | 6.62 | **.003** | 2.27 - 17.12 | + |
|  |  | Recency | .14 | .48 | -.82 - .43 |  |
| **Maternal Adverse Event** | 2.4 | Recency | .34 | **<.001** | .19 - .48 | + |
| **Child Food Insecurity** | 1.4 | Early Childhood | 4.01 | **.002** | 1.48 - 6.28 | + |
| Child Exposure to Community and Domestic Violence | .6 | Middle Childhood | 1.03 | .70 | -42.04 - 14.95 | + |
|  |  | Recency | .14 | .27 | -1.59 - 4.14 |  |
| Maternal IPV | .1 | Recency | .19 | .68 | -1.89 - .33 | - |
| Maternal Substance Abuse | .4 | Accumulation | .91 | .16 | -.96 - 1.71 | + |

Note. The table presents the results from 12 different regression models, one for each adversity category and psychopathology symptom category respectively. All regression models controlled for covariates described previously, with the addition of the EPDS. Coefficients represent unstandardized effect estimates. Bolded values indicate p < .05. Plus (+) symbol indicates at least one common model selected in the case of a combined hypothesis with main analyses. Minus (-) symbol indicates no common model.

Table S5. Sensitivity analysis: Alternative life-course hypothesis operationalization selected by SLCMA for childhood psychopathology symptoms measured via the CBCL

| **Internalizing Symptoms** | | | | | | |
| --- | --- | --- | --- | --- | --- | --- |
| **Adversity Category** | **Partial R-squared %** | **Model Selected** | **Coefficient** | **P-**  **value** | **CI (95%)** | **Concordance with CBCL** |
| **Maternal Psychopathology** | 4.1 | Middle Childhood | 12.23 | **<.001** | 8.67 - 15.41 | + |
| **Maternal Adverse Event** | 1.4 | Recency | .21 | **.003** | .08 - .31 | - |
| **Child Food Insecurity** | 3.2 | Very Early Childhood | -5.86 | **.04** | -11.48 - 1.02 | + |
|  |  | Early Childhood | 7.91 | **.03** | -.56 - 15.28 |  |
|  |  | Recency | .22 | **.03** | -0.01 - .42 |  |
| **Child Exposure to Community and Domestic Violence** | 1.7 | Accumulation | 1.39 | **.001** | .63 - 2.06 | + |
| Maternal IPV | .3 | Accumulation | .54 | .27 | -1.05 - .96 | - |
| Maternal Substance Abuse | .3 | Early Childhood | 2.66 | .22 | -4.11 - 5.06 | - |
| **Externalizing Symptoms** | | | | | | |
| **Adversity Category** | **Partial R-squared %** | **Model Selected** | **Coefficient** | **P-value** | **CI (95%)** | **Concordance with CBCL** |
| **Maternal Psychopathology** | 3.6 | Middle Childhood | 11.87 | **<.001** | 7.84 - 15.86 | + |
| **Maternal Adverse Event** | 1.4 | Recency | .29 | **.002** | .11 - .41 | + |
| Child Food Insecurity | 1.3 | Early Childhood | 8.42 | .06 | -3.0 - 23.79 | + |
|  |  | Recency | .09 | .43 | -.47 - .32 |  |
| Child Exposure to Community and Domestic Violence | 1.6 | Middle Childhood | 1.47 | .52 | -12.57 - 7.78 | + |
|  |  | Recency | .16 | .22 | -.39 - 1.00 |  |
| Maternal IPV | .8 | Very Early Childhood | 1.78 | .47 | -13.90 - 11.79 | + |
|  |  | Accumulation | .44 | .36 | -2.33 - 3.11 |  |
| **Maternal Substance Abuse** | .8 | Very Early Childhood | 4.33 | **.03** | -.11 - 7.36 | - |

Note. The table presents results from 12 separate regression models, one for each adversity category and symptom outcome. All regression models adjusted for covariates. Coefficients represent unstandardized effect estimates. Bolded values indicate p < .05. Plus (+) symbol indicates at least one common model selected in the case of a combined hypothesis with main analyses. Minus (-) symbol indicates no common model.

*BDI*

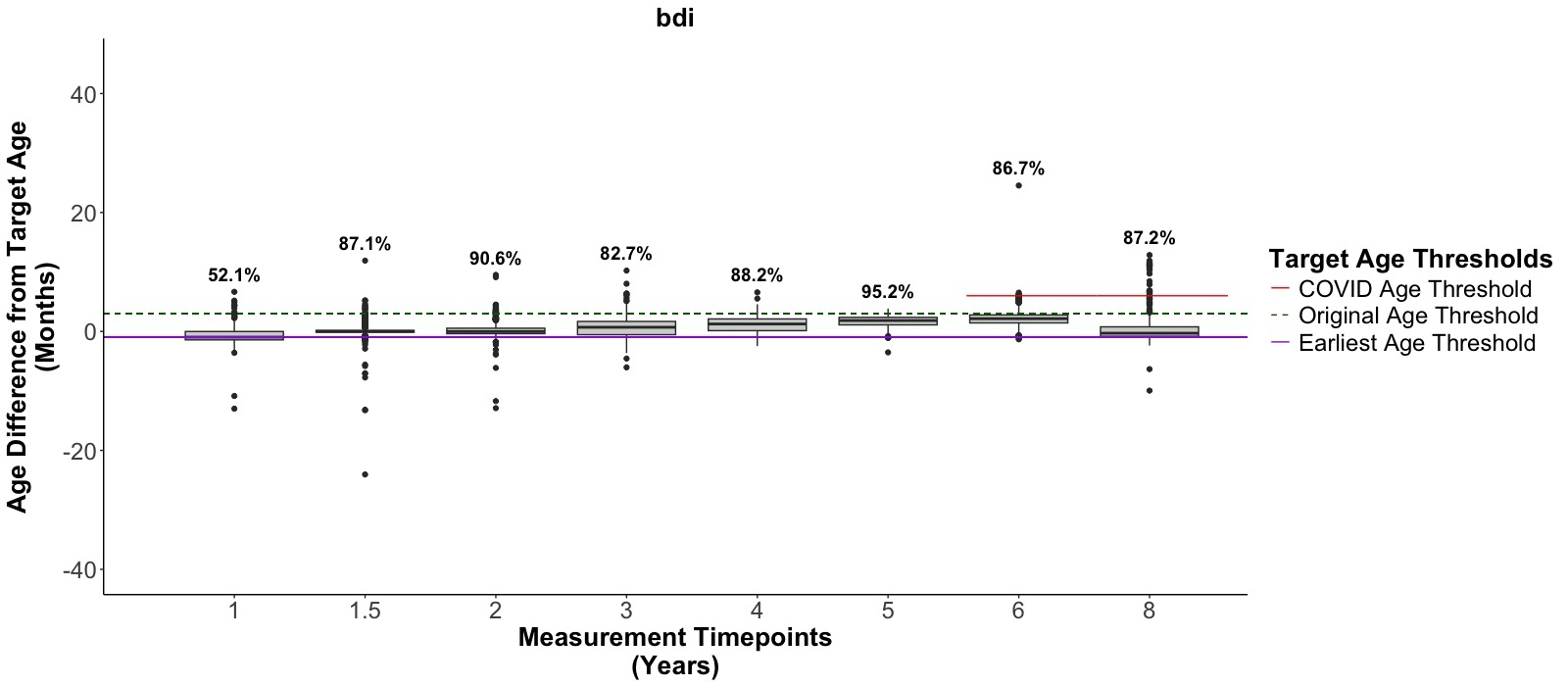

*SRQ*

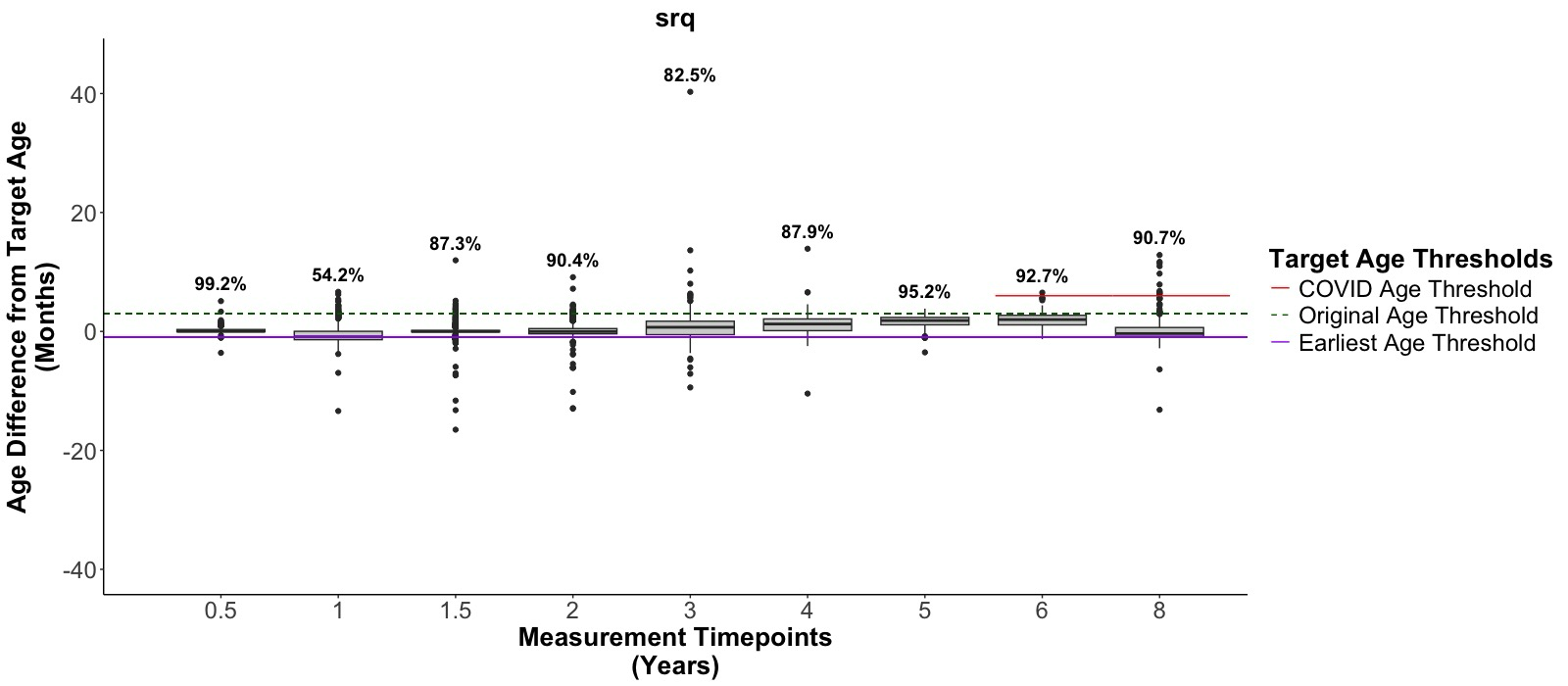

*LEQ*

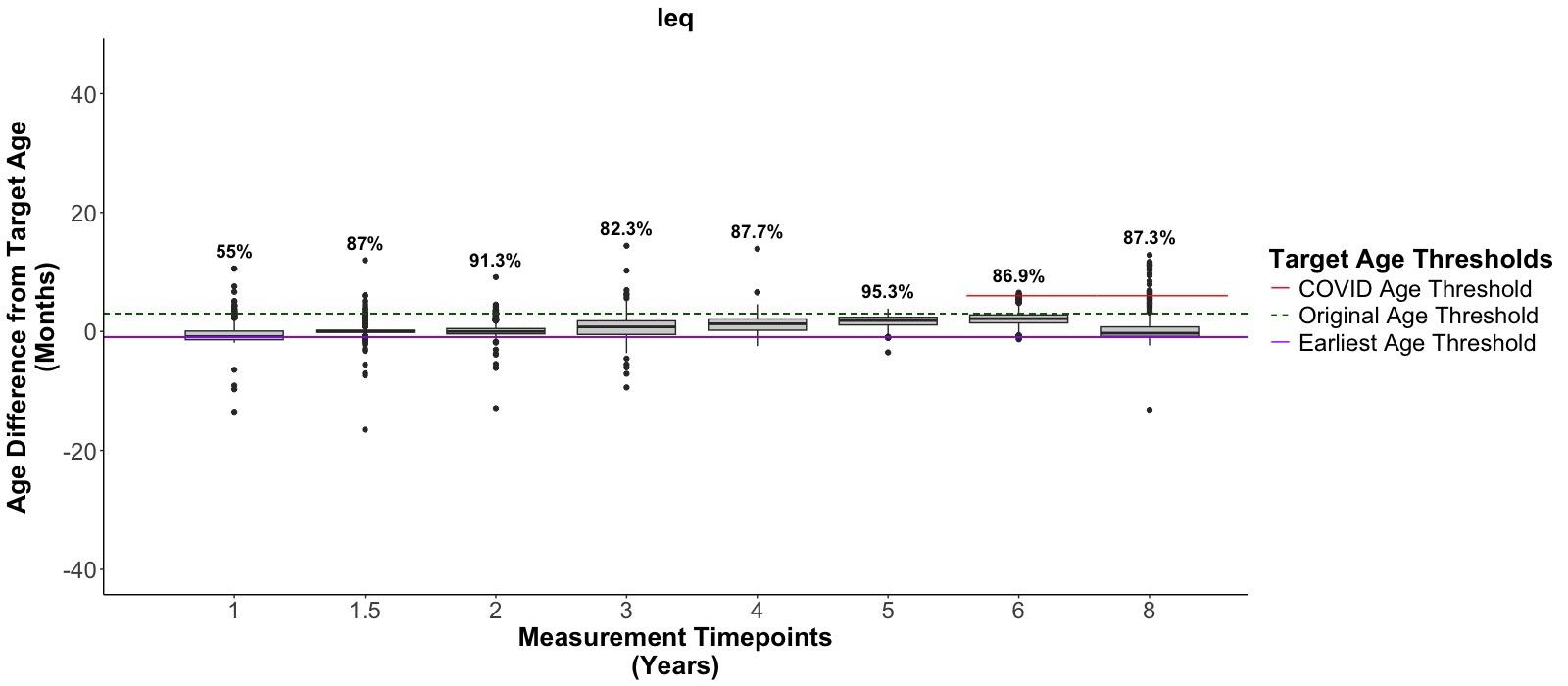

*USDA-HFSS*

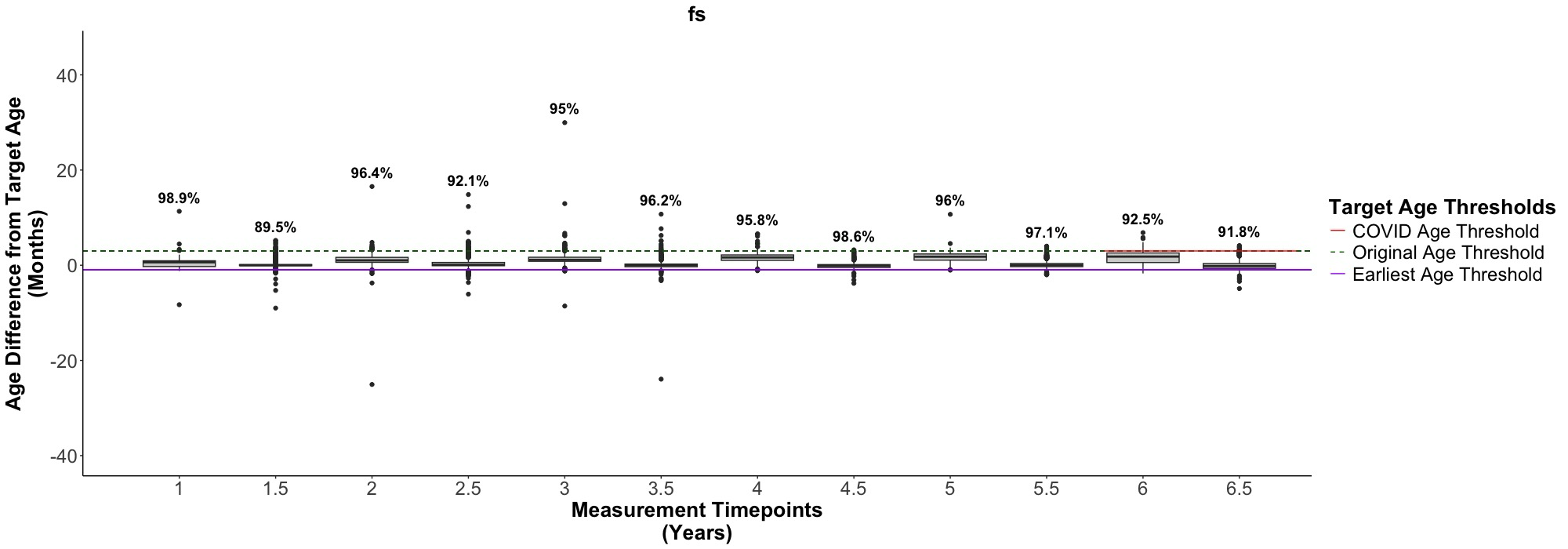

*CECV*

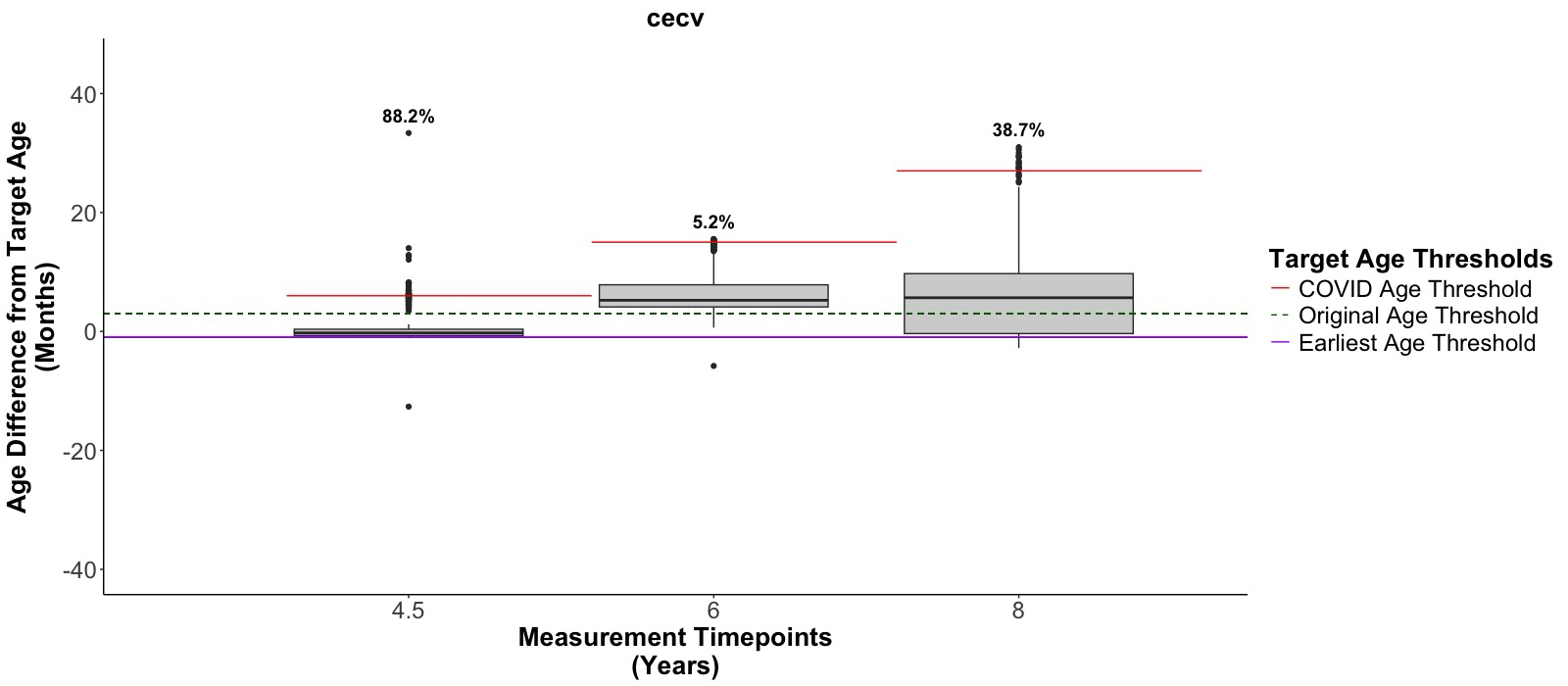

*IPV*

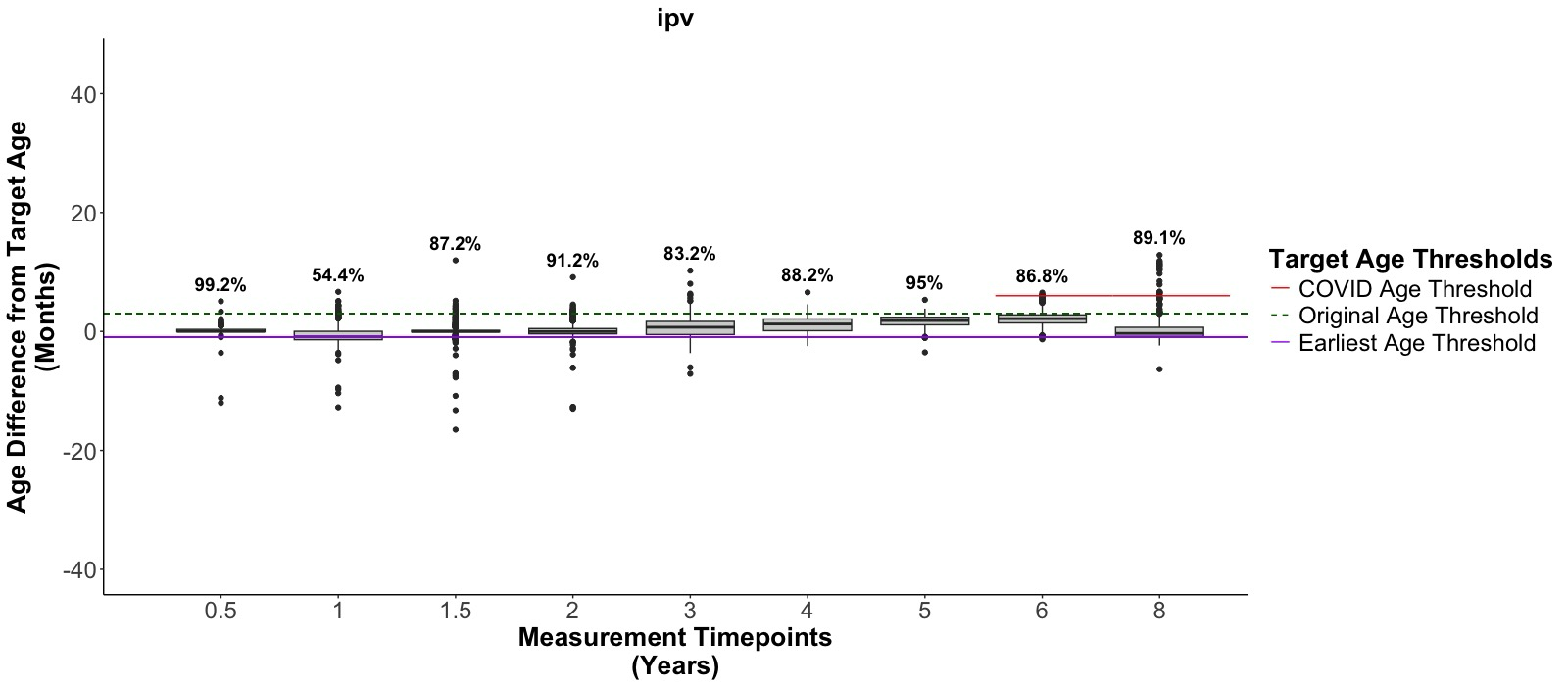

*ASSIST*

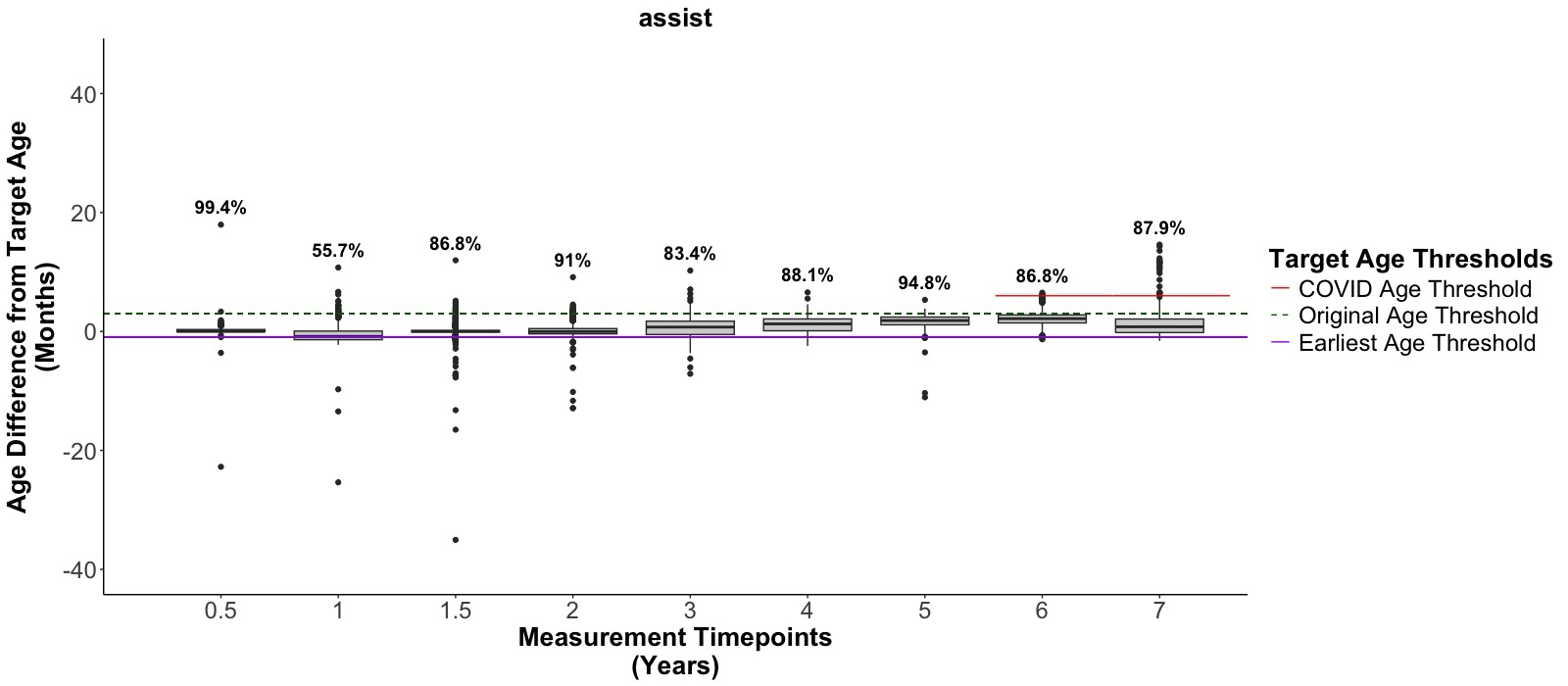

**Figure S1. Percent of data collected within expected age thresholds across time.** Data was expected to be collected within 1 month prior (purple), and 3 months after (green), the child’s birthday. Among the total sample, we report the percent of data that meets the criteria of falling between the threshold (between purple and green lines) among data that was successfully collected at each timepoint. The threshold was expanded to 6 months after the birthday due to the COVID-19 pandemic, with the exception of the USDA-HFSS and CECV (red line). Although data was collected at high fidelity during the COVID-19 pandemic within the original threshold, it increased by 1.9% to 92.7% when incorporating the expanded range. BDI = Beck Depression Inventory. SRQ = Self-Reporting Questionnaire. LEQ = Life Events Questionnaire. USDA-HFSS = US Department of Agriculture Short Form Household Food Security Scale. CECV = Child Exposure to Violence Checklist. IPV = Intimate Partner Violence Questionnaire. ASSIST = Alcohol, Smoking, and Substance Involvement Screening Test.

*BDI*

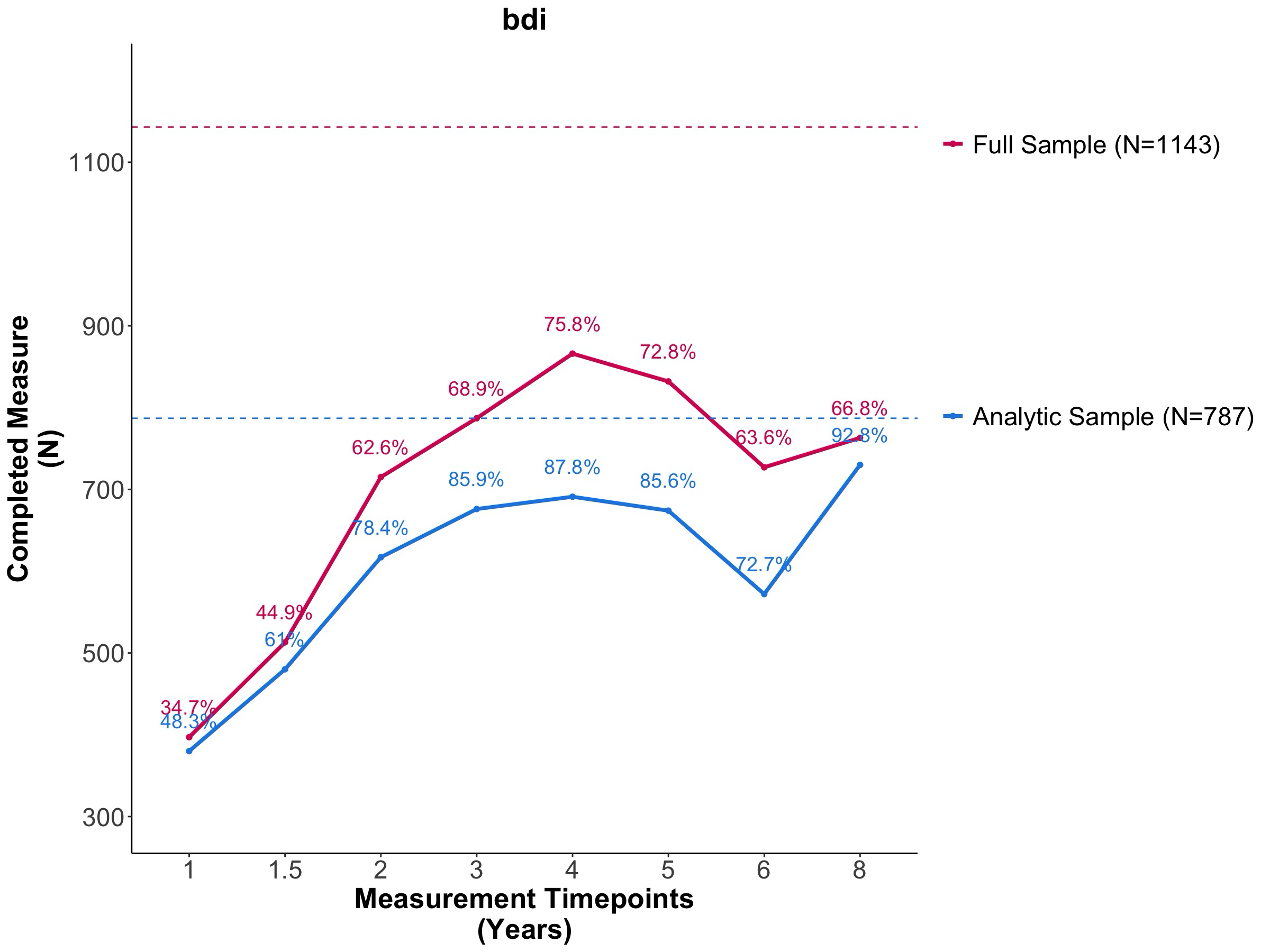

*SRQ*

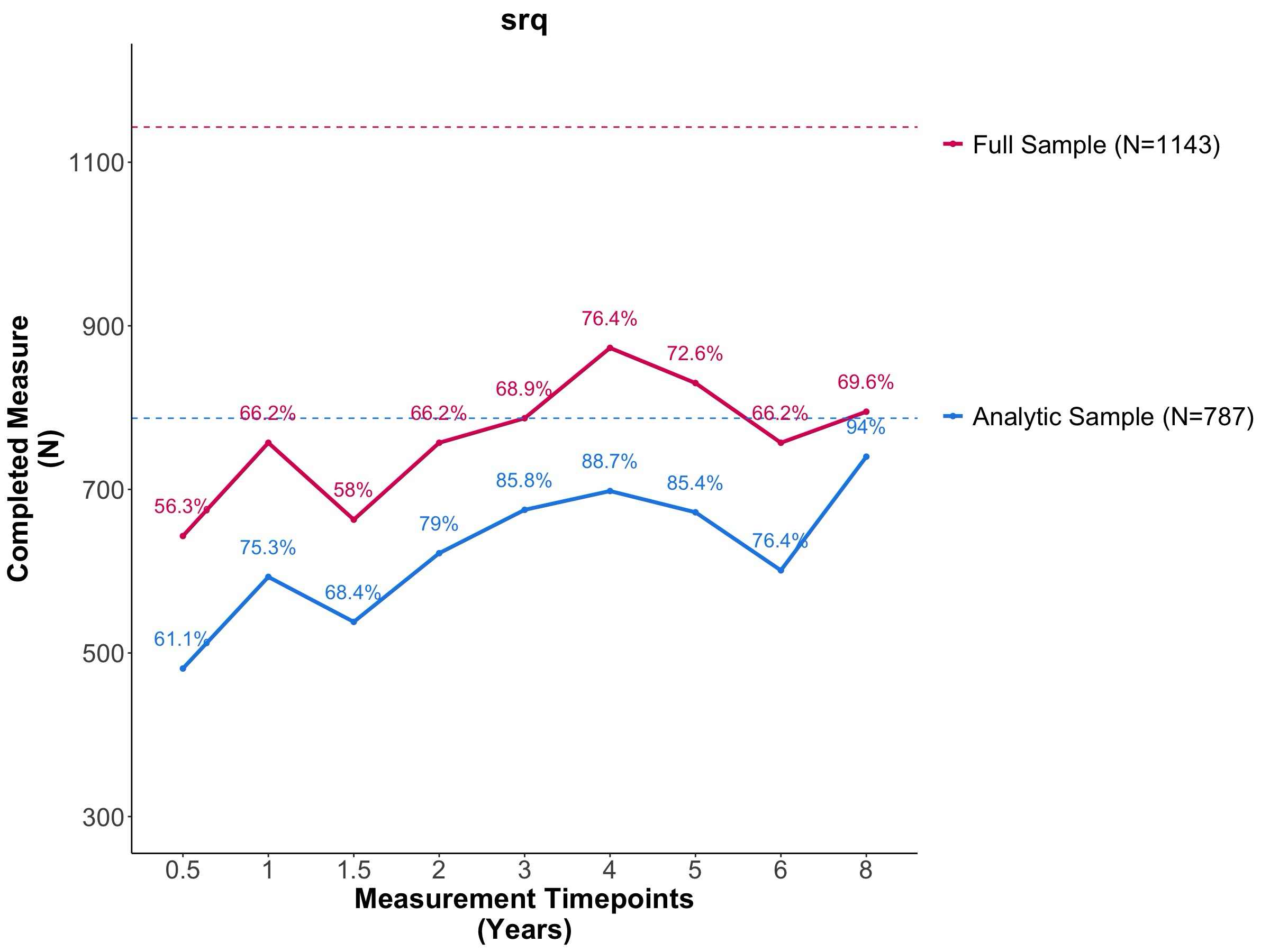

*LEQ*

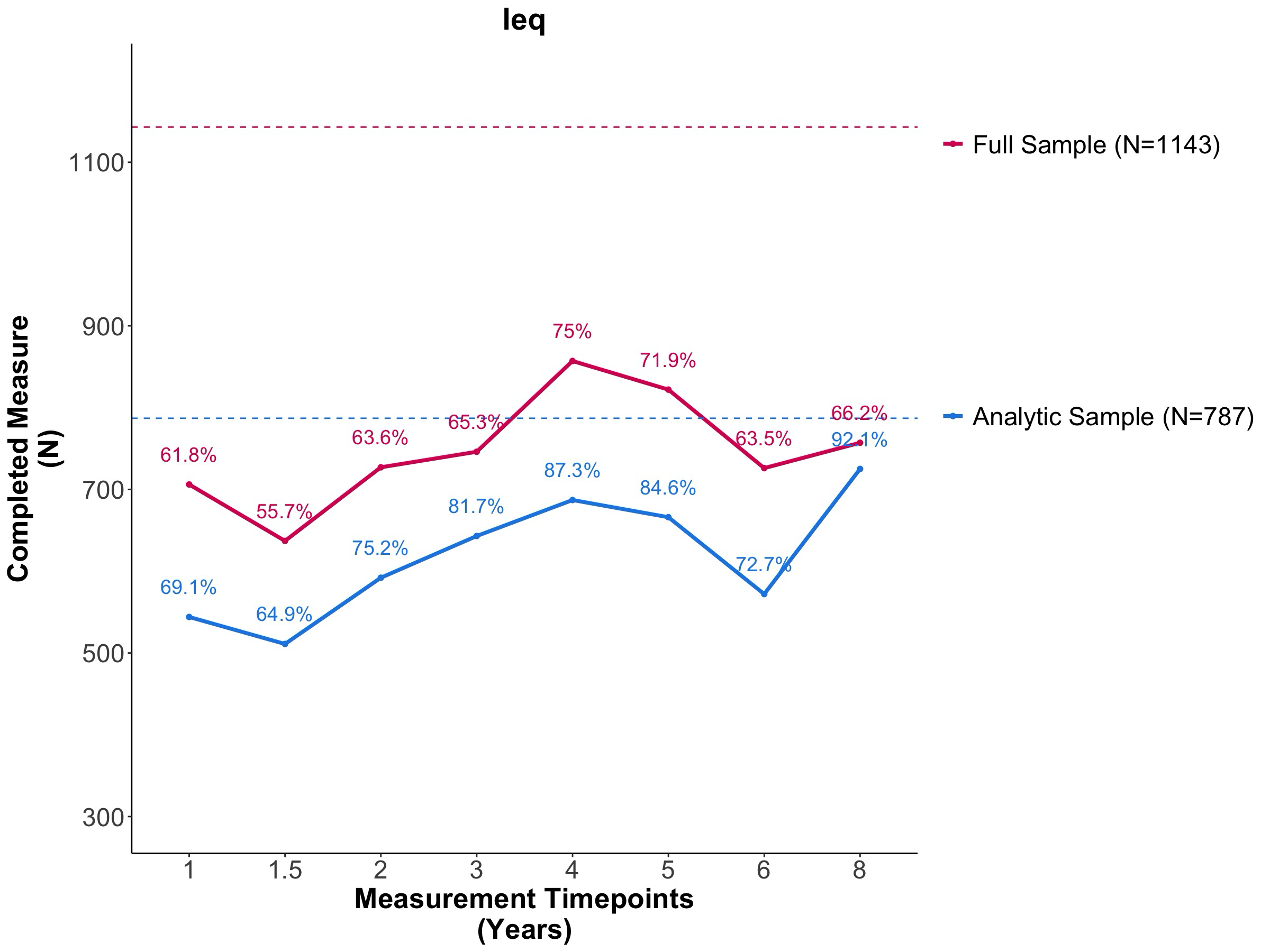

*USDA-HFSS*

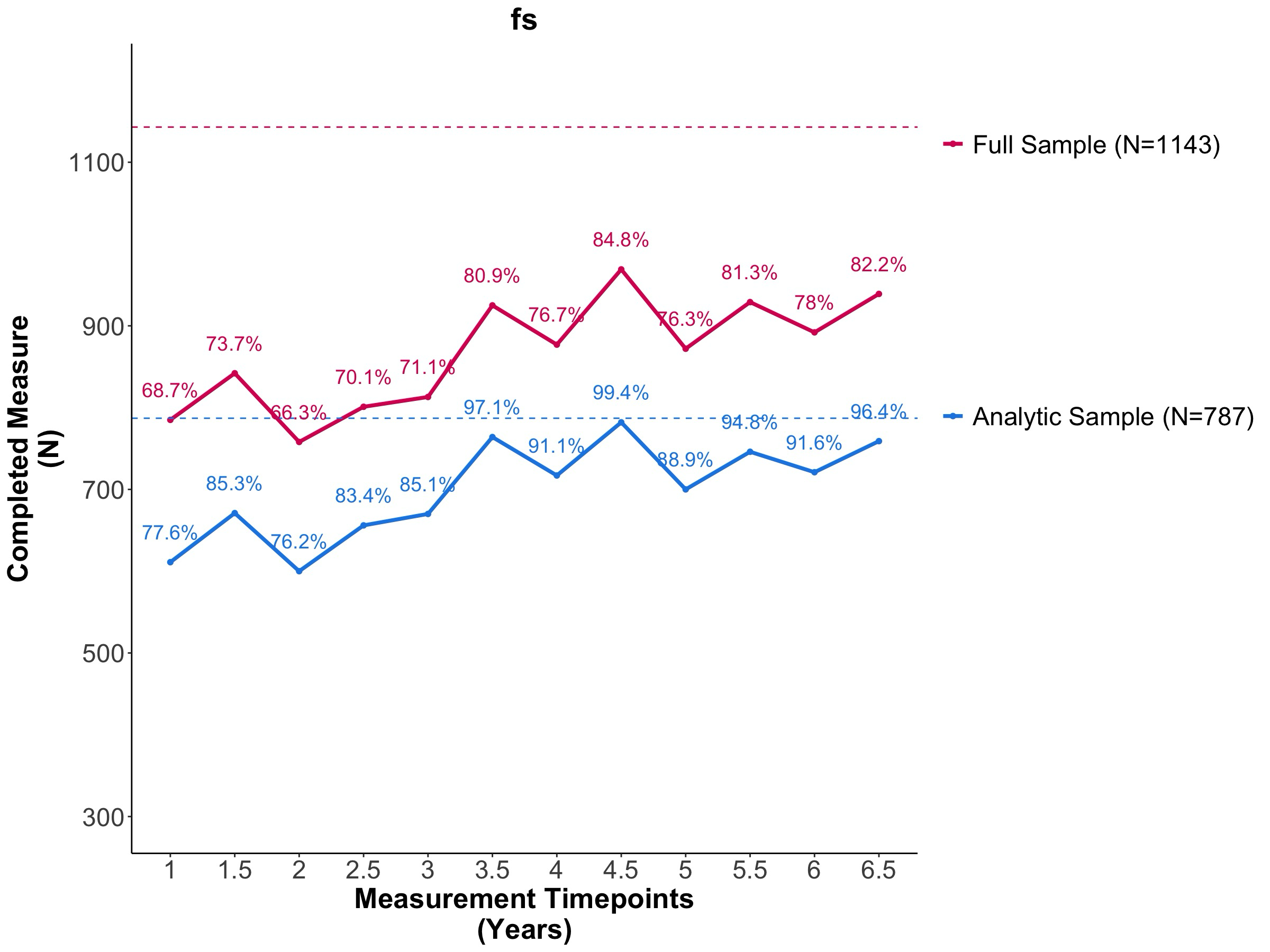

*CECV*

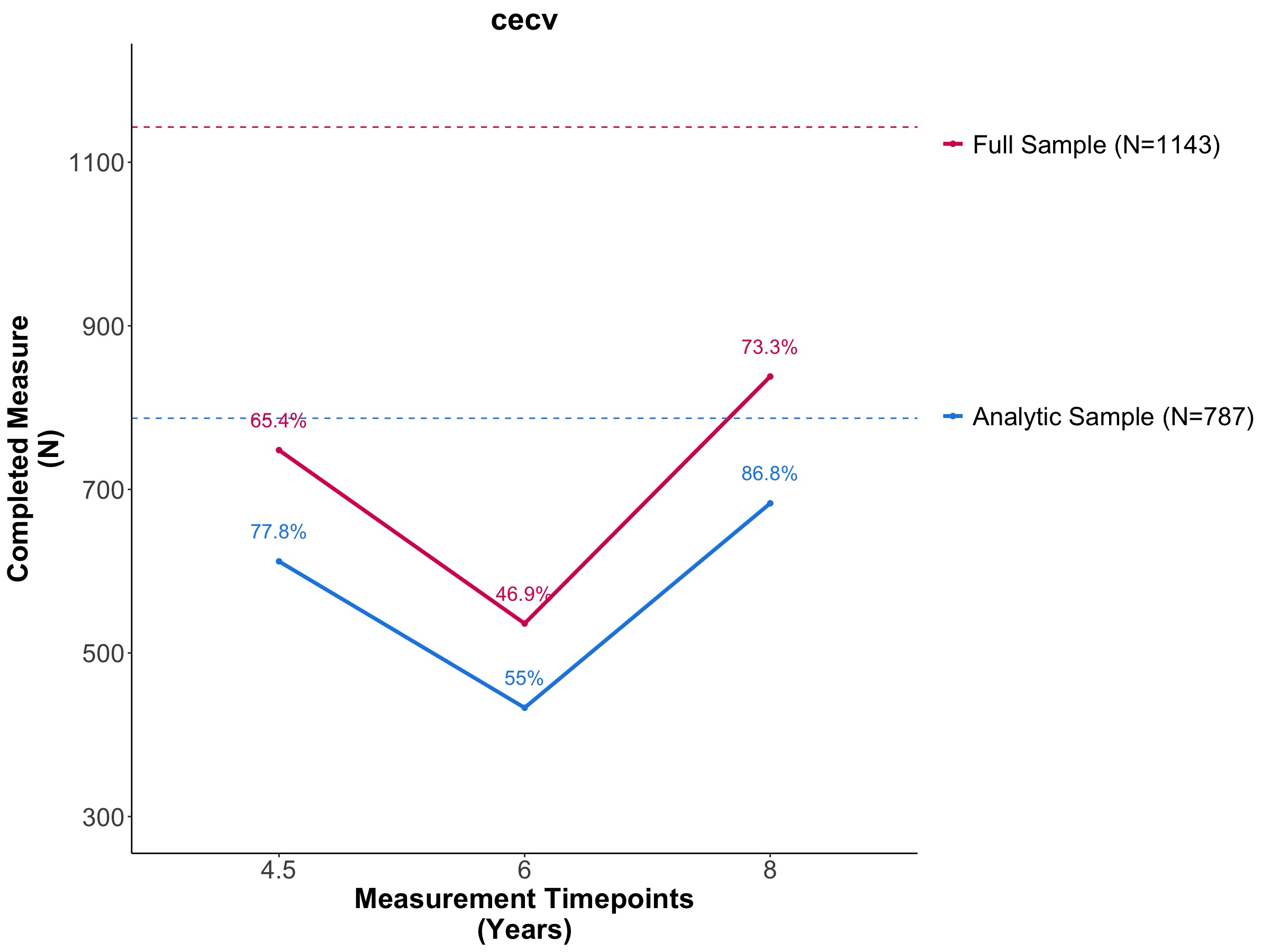

*IPV*

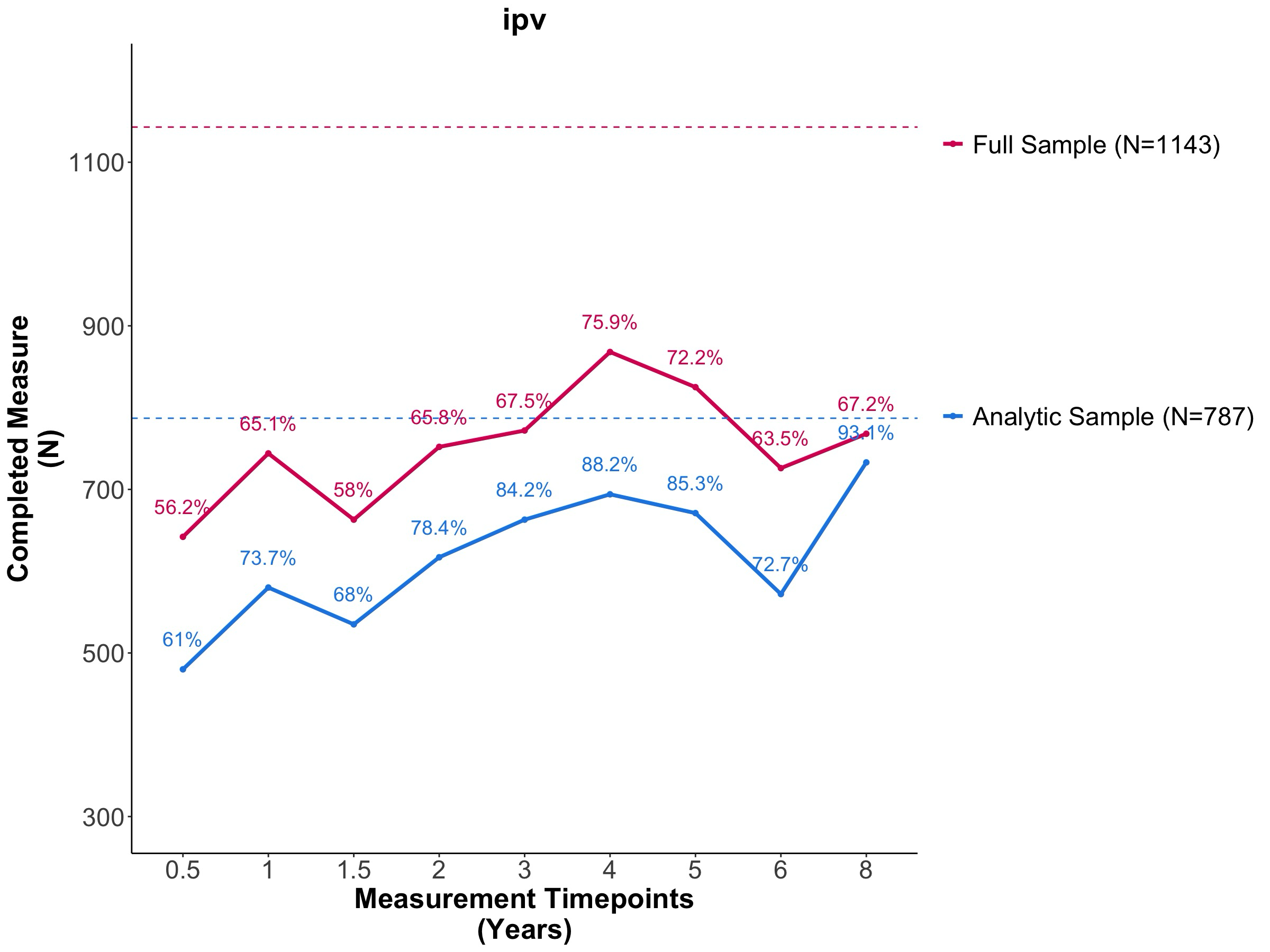

*ASSIST*

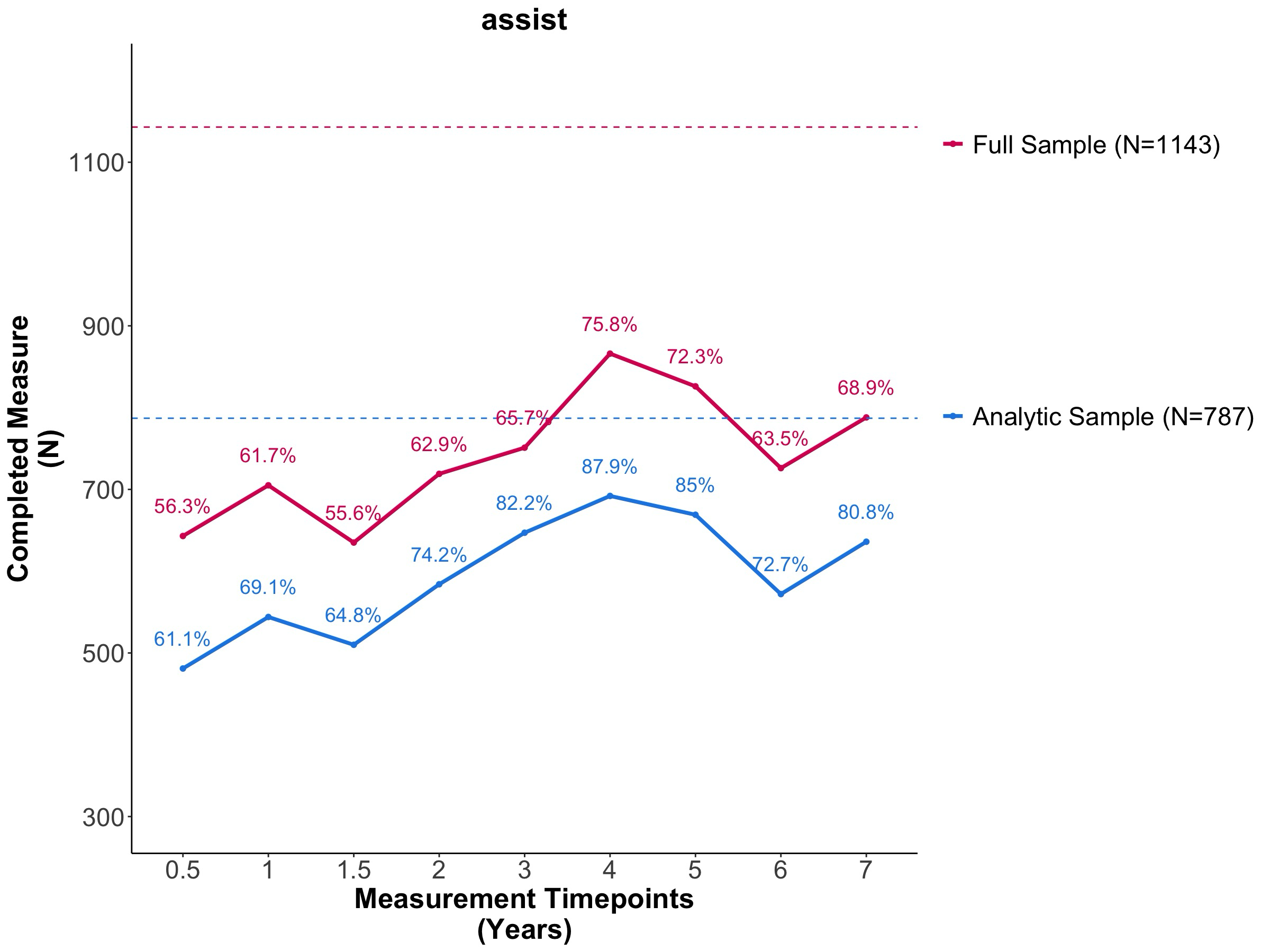

**Figure S2. Percent data collected at each timepoint.** Lineplots show the amount of non-missing data collected across timepoint for each measure for both the analytic sample (blue) and full sample (red). BDI = Beck Depression Inventory. SRQ = Self-Reporting Questionnaire. LEQ = Life Events Questionnaire. USDA-HFSS = US Department of Agriculture Short Form Household Food Security Scale. CECV = Child Exposure to Violence Checklist. IPV = Intimate Partner Violence Questionnaire. ASSIST = Alcohol, Smoking, and Substance Involvement Screening Test.

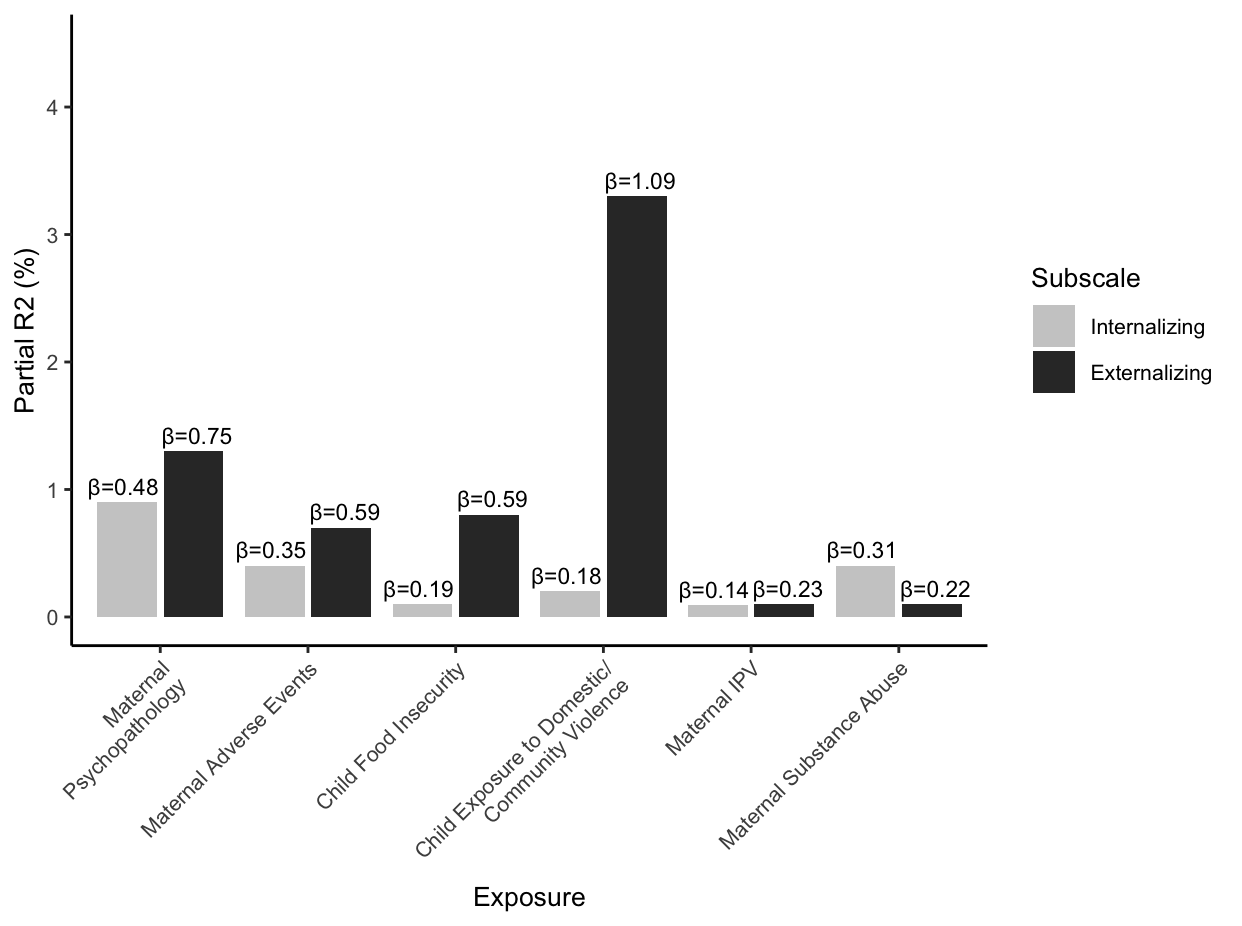

**Figure S3. Associations of exposure to each type of adversity with SDQ-measured psychopathology symptoms.** Bars display variance explained (*R^2^*) by each adversity. Values displayed above each bar depict unstandardized effect estimates. Results were obtained from ever-exposed – i.e., irrespective of life-course hypothesis selection – multiple regression analyses, adjusted for covariates. IPV = intimate partner violence. SDQ = Strengths and Difficulties Questionnaire.

**CBCL**

*Internalizing*

A) Maternal Psychopathology	B) Maternal Adverse Event

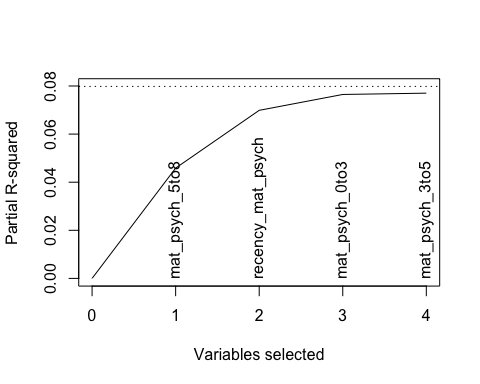

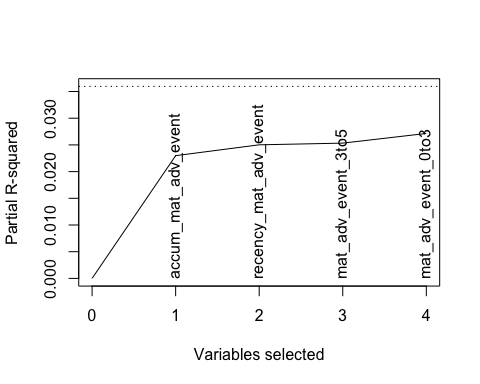

C) Child Food Insecurity	D) Child Exposure to Domestic/Community Violence

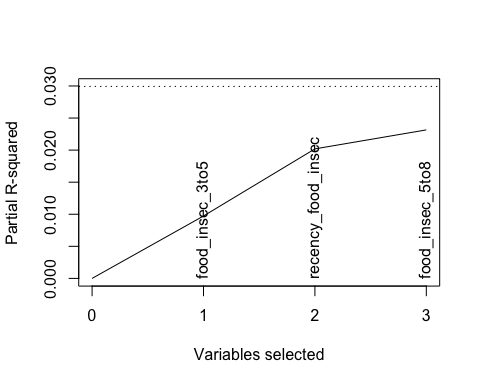

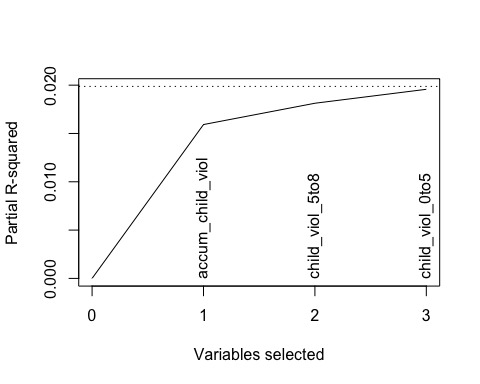

E) Maternal Intimate Partner Violence	F) Maternal Substance Abuse

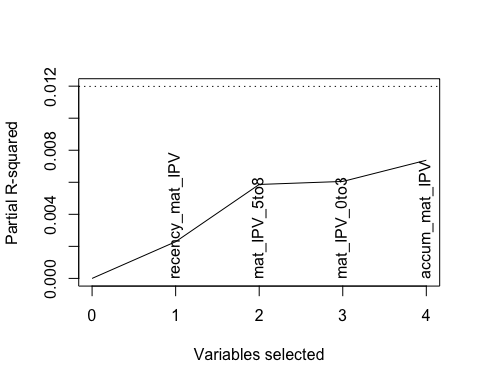

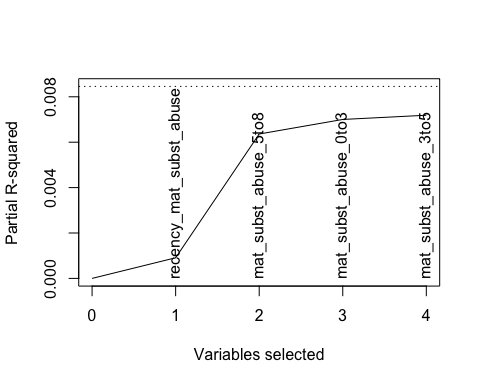

*Externalizing*

A) Maternal Psychopathology	B) Maternal Adverse Event

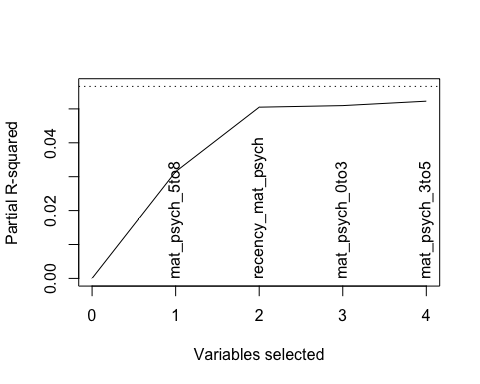

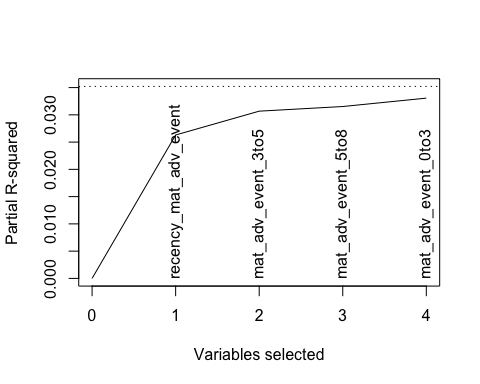

C) Child Food Insecurity	D) Child Exposure to Domestic/Community Violence

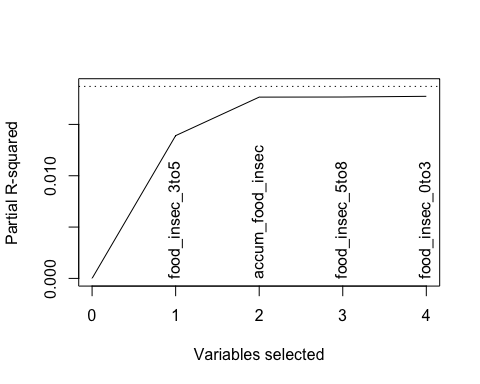

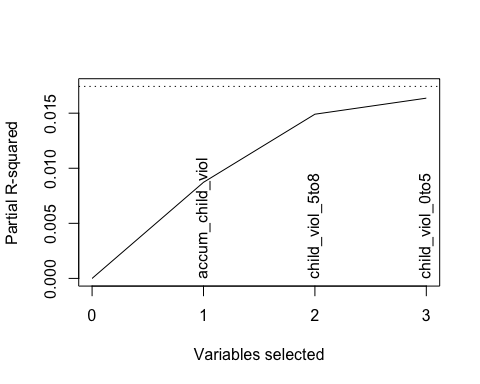

E) Maternal Intimate Partner Violence	F) Maternal Substance Abuse

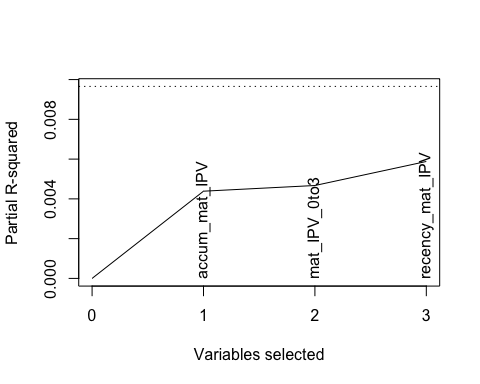

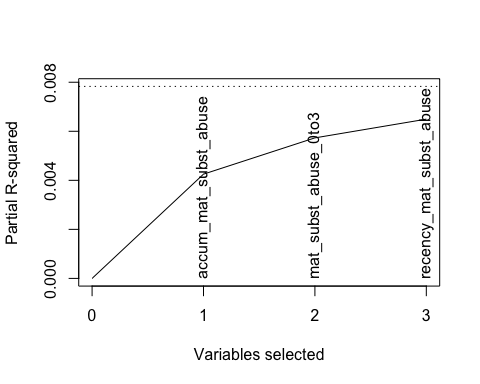

**Figure S4. Elbow plots illustrating LARS variable selection procedure.** Elbow plots depict the results of the LARS selection procedure for the effects of each adversity model on internalizing or externalizing symptoms. The procedure tests sensitive period, recency, and accumulation hypotheses. LARS begins by identifying which individual variable has the strongest association to the outcome; it then identifies the combination of two variables with the strongest association, and so on. As a result, the plot shows the overall increase in R2 in the model as additional predictors are added. The point where each plot begins to flatten (or the “elbow”) indicates the point where there are diminishing returns to the model goodness-of-fit. Thus, the variables selected prior to the elbow optimize the least amount of variables that have the greatest effect. LARS = least angle regression.

**CBCL**

*Internalizing*

A) Maternal Psychopathology	B) Maternal Adverse Event

Middle Childhood + Recency	Accumulation

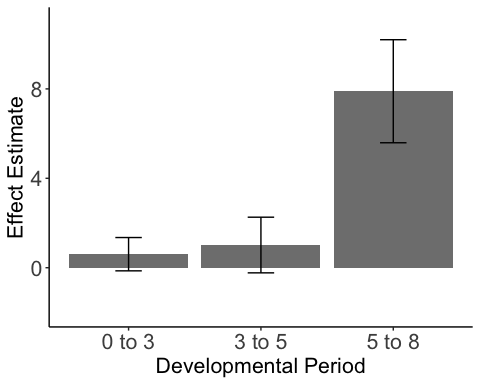

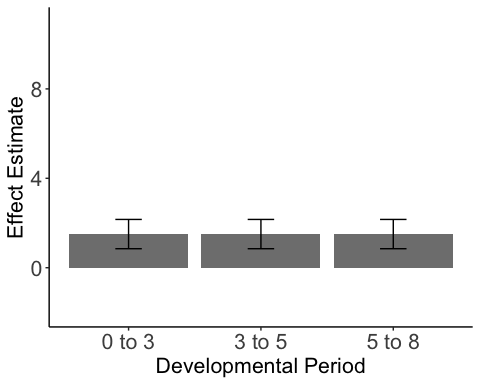

C) Child Food Insecurity	D) Child Exposure to Domestic/Community	Violence

Early Childhood + Recency	Accumulation

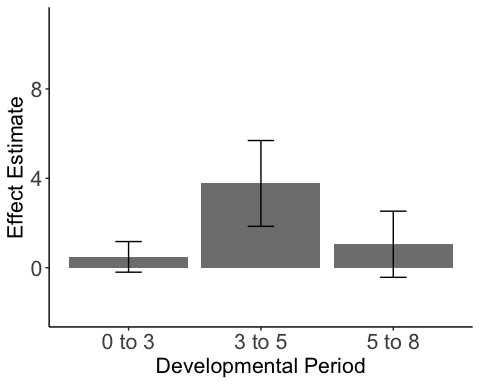

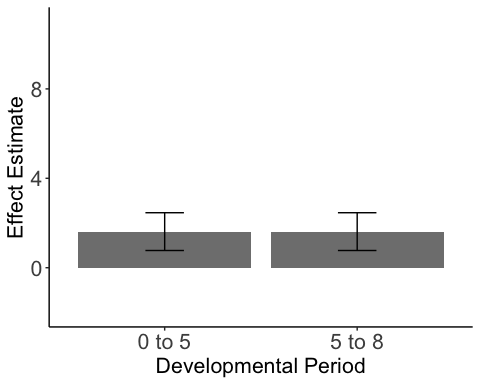

E) Maternal Intimate Partner Violence	F) Maternal Substance Abuse

Middle Childhood + Recency	Middle Childhood + Recency

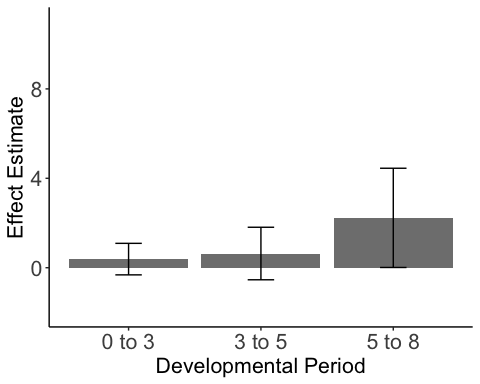

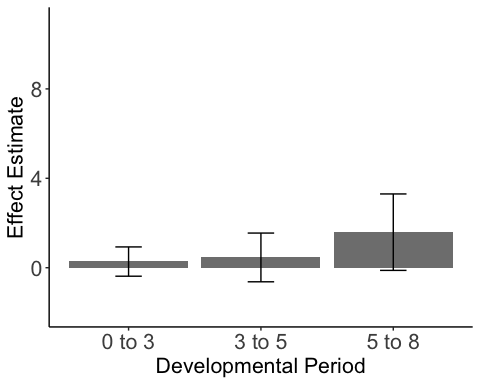

*Externalizing*

A) Maternal Psychopathology	B) Maternal Adverse Event

Middle Childhood + Recency	Recency

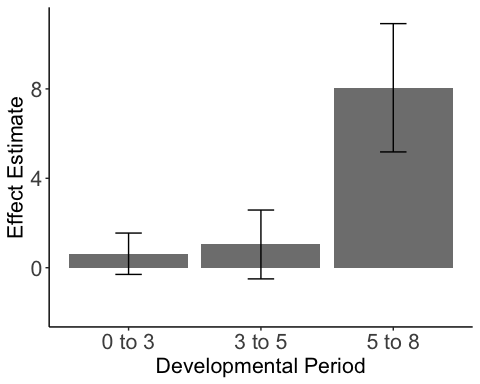

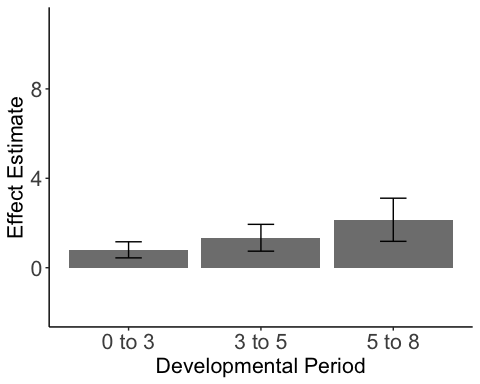

C) Child Food Insecurity	D) Child Exposure to Domestic/Community	Violence

Early Childhood	Middle Childhood + Accumulation

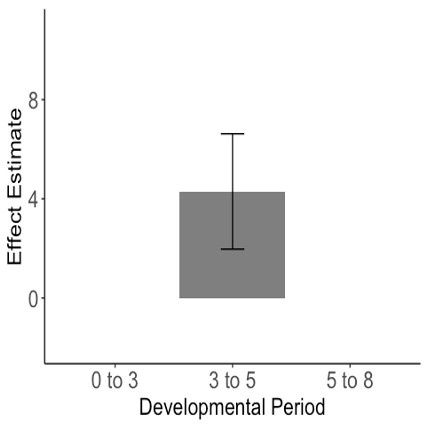

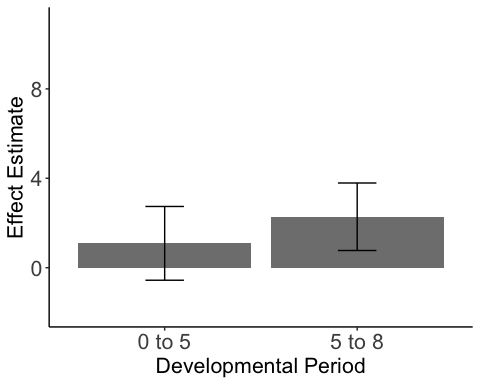

E) Maternal Intimate Partner Violence	F) Maternal Substance Abuse

Accumulation	Accumulation

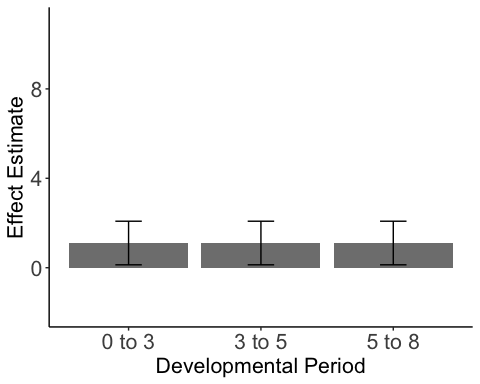

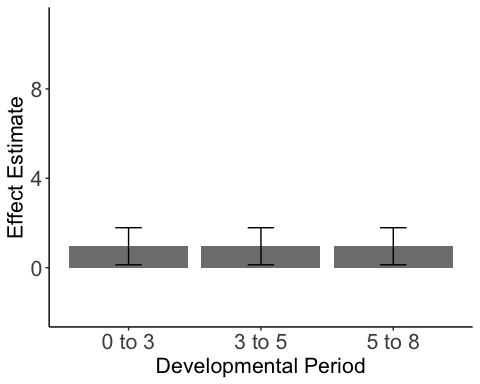

**Figure S5. Effects of adversity exposure during each developmental period, based on the selected SLCMA life-course hypotheses via the CBCL for all adversity categories.** Figures illustrate the weighted effect of exposure during each developmental period based on the life-course hypothesis/es selected for each adversity. SLCMA = Structured Life-course Modeling Approach. CBCL = Child Behavior Checklist.
